# Discontinuation of antihypertensive and lipid-lowering medication in primary care: a systematic review of observational data and socio-demographic differences

**DOI:** 10.64898/2026.04.28.26351691

**Authors:** Simon R. Parker, Nalin Natarajan, Cini Bhanu, A Floriaan Schmidt, Nishi Chaturvedi, Sophie V. Eastwood

**Affiliations:** Unit for Lifelong Health and Ageing, Department of Population Science and Experimental Medicine, Institute of Cardiovascular Science, Faculty of Population Health, University College London, London, UK; BHF Centre of Research Excellence, University College London, London, UK; Department of Primary Care and Population Health, University College London, London, UK; Department of Cardiology, Amsterdam Cardiovascular Sciences, Amsterdam University Medical Centre, University of Amsterdam, Amsterdam, The Netherlands

## Abstract

**Background:** Cardiovascular disease (CVD) risk is chiefly managed pharmacologically in primary care using lipid-lowering therapies (LLTs) and antihypertensives (AHTs), but patients frequently discontinue treatment. Little up-to-date synthesised real-world data for LLT or AHT discontinuation, or how it varies by socio-demographic group, exists.

**Methods:** We systematically reviewed observational studies from PubMed, EMBASE, Web of Science, and CINAHL from 2010-2025 describing discontinuation/restarting prevalence for first-to-third line LLTs/AHTs used for CVD prevention in primary care. Data were extracted on discontinuation/restarting prevalences and associations between discontinuation and sociodemographic factors.

**Results:** Of 5,756 records, 44 (29 LLT; 15 AHT) reports were included. LLT median (IQR) discontinuation and restarting prevalences were 43% (33%-59%) and 58% (40%-61%), respectively. AHT discontinuation and restarting prevalences were 41% (30%-49%) and 28%, respectively. Discontinuation/restarting prevalence depended on discontinuation definition and indication.

Patients aged ≈65 years were less likely to discontinue than younger or older patients, for both LLTs and AHTs. In general, women appeared to discontinue LLTs more often than men; men discontinued AHTs more often for primary prevention. Income-based socioeconomic position (SEP) measures were associated with discontinuation, but composite SEP measures were not. Minority ethnic groups were more likely to discontinue LLTs and AHTs.

**Conclusions:** This systematic review of real-world data identified high levels of LLT and AHT discontinuation, and inequalities according to age, sex, and ethnicity. Awareness of these patterns and additional research into patient-level drivers of drug discontinuation could improve health equity by addressing LLT/AHT discontinuation in the highest-risk patients.

## INTRODUCTION

Cardiovascular disease (CVD) is the leading cause of mortality and morbidity worldwide.^1^ Clinical trials show that antihypertensive (AHTs) and lipid-lowering therapies (LLTs) can reduce CVD by up to 30% in both primary^2,3^ and secondary^4,5^ prevention populations. However, frequent medication discontinuation may preclude these benefits in practice.^6^

CVD incidence increases with age and varies starkly by sociodemographic group. Incidence of most CVDs are two- to five-fold greater for men than women,^7^ 20% higher for those living in less versus more socio-economically disadvantaged circumstances^8^ and up to 50% higher in some ethnic groups.^9^ Importantly, discontinuation of CVD prevention medication may also vary by sociodemographic group; both younger (<50 years old) and/or older (>70 years old) groups, females, and individuals with higher incomes may be less likely to discontinue statins, though these patterns are not consistent across studies.^10–12^ Moreover, our previous work indicates South Asian and African/African-Caribbean AHT users may be more likely to discontinue in the United Kingdom compared to White European individuals.^13^ These differences in discontinuation may correspond with and contribute to inequalities in CVD outcomes.

Therefore, accurate ascertainment of overall and group differences in CVD preventative medication discontinuation is crucial for targeting healthcare. This data must be gleaned from “real-world” contexts, as opposed to trials, where strict inclusion criteria and rigorous follow-up may falsely reduce discontinuation rates and limit generalisability to whole population use. Also, discontinuation should be assessed in primary care, given that primary care contexts account for the most use of LLTs and AHTs.

Previous reviews have addressed statin^10–12,14^ or antihypertensive^15–18^ use and included socio-demographic group differences, but findings are limited by their vintage (most are ≥10 years old), inclusion of trial and observational data (despite reduced generalisability of the former),^10–12,14,18^ focus on adherence rather than discontinuation (despite the latter having greater implications for CVD risk),^16,18^ absence of data on ethnicity or socio-economic differences,^10,11,16^ and lack of data on switching or re-starting treatments.^11,12,14,16,18^ Additionally, no study that we are aware of compares aggregated data on patterns in AHT and LLT discontinuation simultaneously, though examining usage patterns of the two cornerstones of primary care preventative treatment together may generate insights into both drivers and implications of discontinuation.

Therefore, we aimed to 1) provide a systematic, up-to-date evidence synthesis of observational data on discontinuation of AHTs and LLTs in primary care, and, where possible, 2) describe discontinuation patterns by age, sex, socio-economic position (SEP) and ethnicity, 3) assess whether discontinuation patterns vary by primary/ secondary prevention and 4) describe AHT and LLT restarting rates.

## METHODS

We performed a systematic review with a pre-specified study protocol (PROSPERO ID: CRD420250599340), though limited deviations from this protocol were made (online supplemental file, p2). This study is reported per the Systematic reviews and Meta-Analyses (PRISMA) guidelines (online supplemental file, p7).^19^

### Search strategy

We searched PubMed, EMBASE, Web of Science, and CINAHL. The search strategy utilised free-text and subject keywords (e.g. MeSH for PubMed) for antihypertensives (AHTs) and lipid-lowering therapies (LLTs), combined with free-text terms for medication adherence, discontinuation, and deprescribing (online supplemental file, p2). We excluded randomised controlled trial (RCT) data using the Cochrane collaboration’s recommended filter.^20^ Searches were restricted to after 2010 to better reflect current clinical practice.^21,22^

### Inclusion criteria

Included studies were required to use electronic health records (EHRs) to assess discontinuation rates in adults (≥18 years old) initiating first- to third-line lipid- or blood pressure-lowering medications in primary care (online supplemental file; p6). For specific drug classes within each category, see supplementary methods table (online supplemental file; p6). For the purposes of this review, EHRs were any large-scale electronic system with digitised prescribing and/or dispensing records. Our procedure for papers with multiple drug classes is included in the supplementary methods table (online supplemental file; p6). Included studies were required to use primary care EHRs.

### Study selection

Two authors (SP and NN) independently screened abstracts, with SVE acting as a third independent arbiter. SP accessed full-text articles for rescreening against the eligibility criteria. Papers with unclear eligibility at this stage were referred to another author (SVE) for discussion on inclusion.

### Data extraction

Summary data were extracted for each included article including study characteristics, results from both unadjusted and adjusted regression models and, where available, therapy restarting incidence. Where data in a paper was presented across sub-cohorts, we combined data from these sub-cohorts into overall paper-level cohorts, using the Cochrane formulae for combining groups (online supplemental file, p5).^23^ Where studies reported discontinuation prevalences from multiple methodologies, we collected all measures and defined them by the discontinuation description in use. Where reported, we collected counts of participants discontinuing/remaining on treatment within each age/sex/SEP/ethnic group.

### Assessment of study quality

Studies were assessed for quality by SP using the Risk Of Bias In Non-randomized Studies - of Exposures (ROBINS-E) tool.^24^

### Statistical analysis

All data visualisation and statistical analyses were performed using R (v. 4.4.1).

For papers which reported discontinuation counts for participant categories of interest (age, sex, SEP, ethnicity), we estimated risk ratios for discontinuation using logistic models, with standard errors calculated using the conditional standardisation method using the *risks* package.^25,26^ These were combined with results from adjusted regression analyses (where performed). Where necessary, reference categories were harmonised across studies (online supplemental file, p6). Meta-analysis of effect estimates for association between sociodemographic group and discontinuation was precluded by study heterogeneity. Data were visualised using forest plots.

### Patient and public involvement

Patients or the public were not involved in the design, conduct, reporting, or dissemination plans of this research.

## RESULTS

5,756 unique records were retrieved from PubMed, Web of Science, EMBASE, and CINAHL Plus. Of 482 records selected for full text review, 31 were selected for inclusion (Figure 1); 16 with data for LLTs, and 15 with data for AHTs. 13 additional articles for LLTs were found in the references of relevant systematic reviews,^11,12,14,27,28^ but none from the references of included papers.

**Figure 1.**
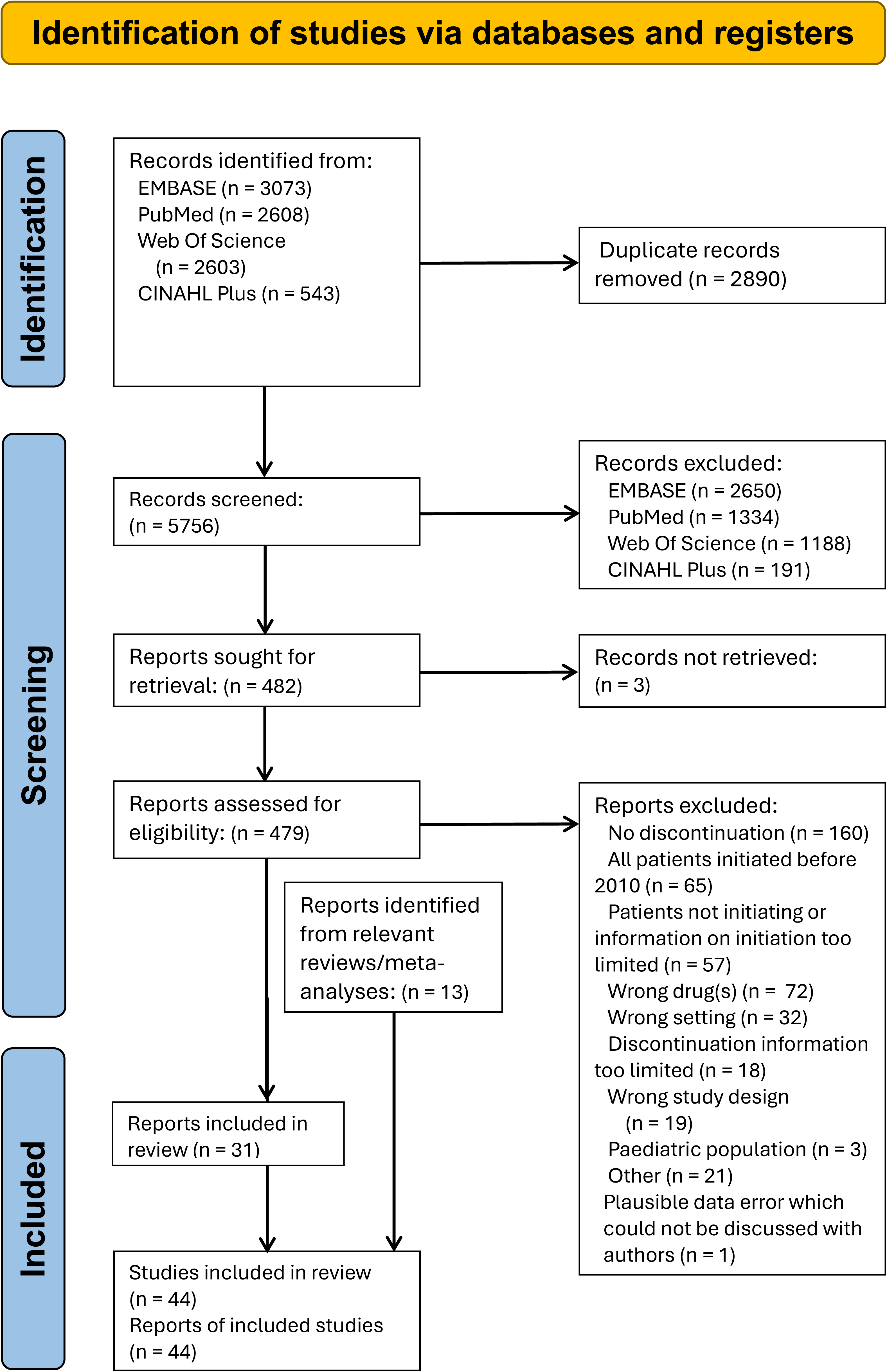
PRISMA flow chart diagram.

The included papers had data from 11,645,049 patients, with database re-use across the included papers (Table 1). Mean age ranged from 51^29^ to 80^30^ years, and the percentage of males from 31%^31^ to 100%^32^. Of fifteen AHT papers, three were based in Asia, eight in Europe, three in North America, and one in Oceania (Australia only). Of 28 LLT papers, six were based in Asia, eleven in Europe, three in North America (United States only), and six in Oceania.

**Table 1.**
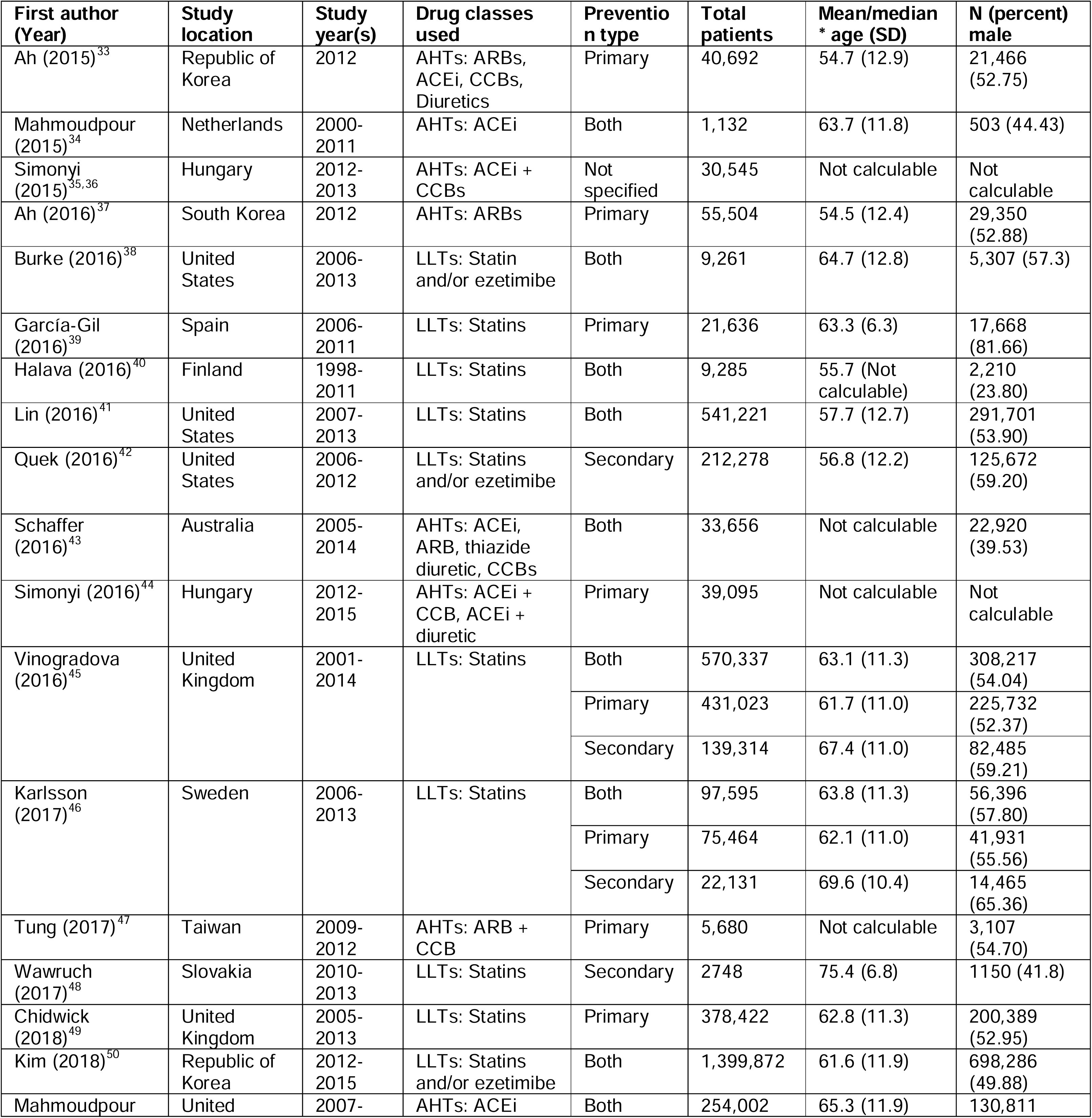
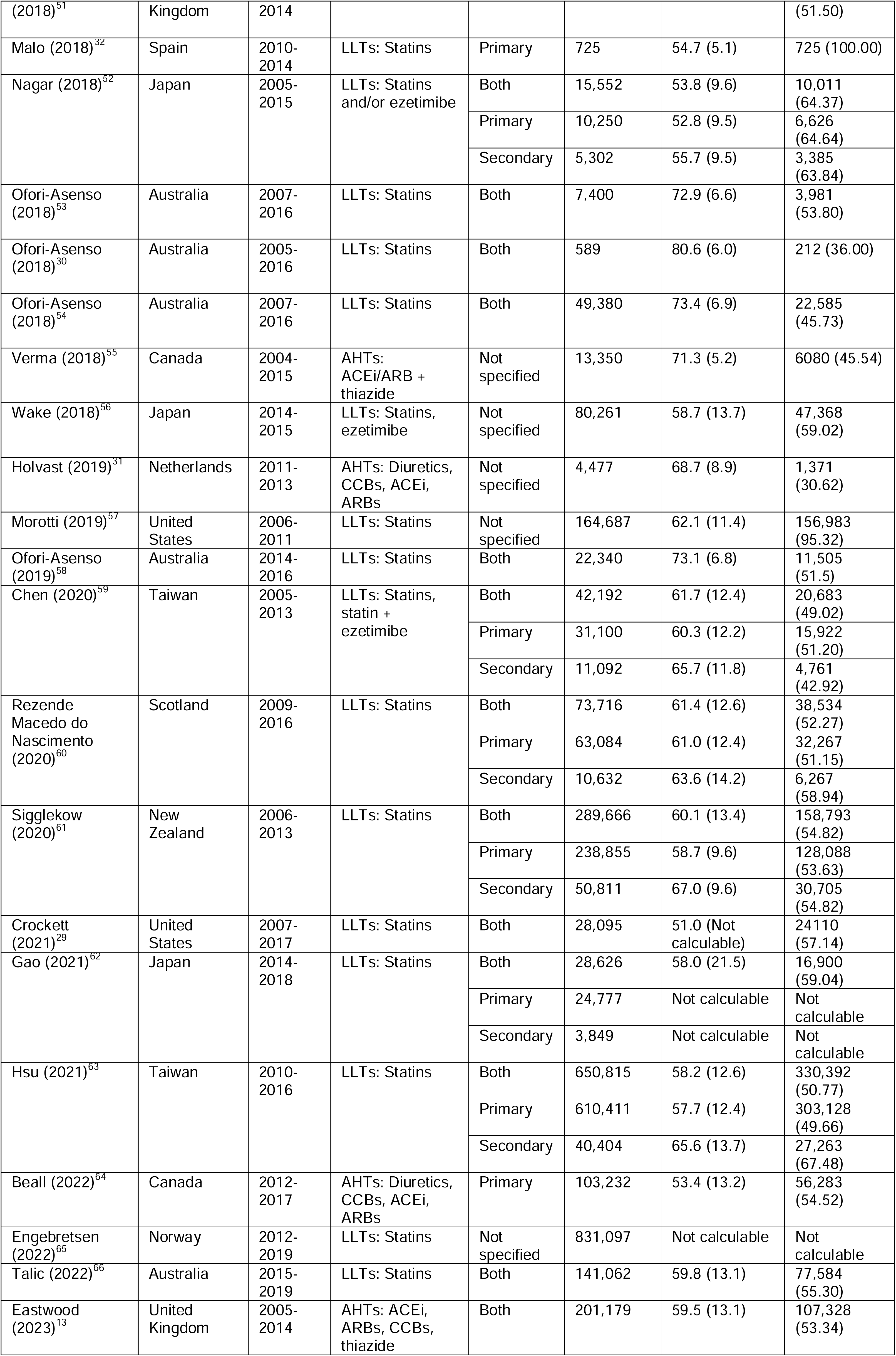
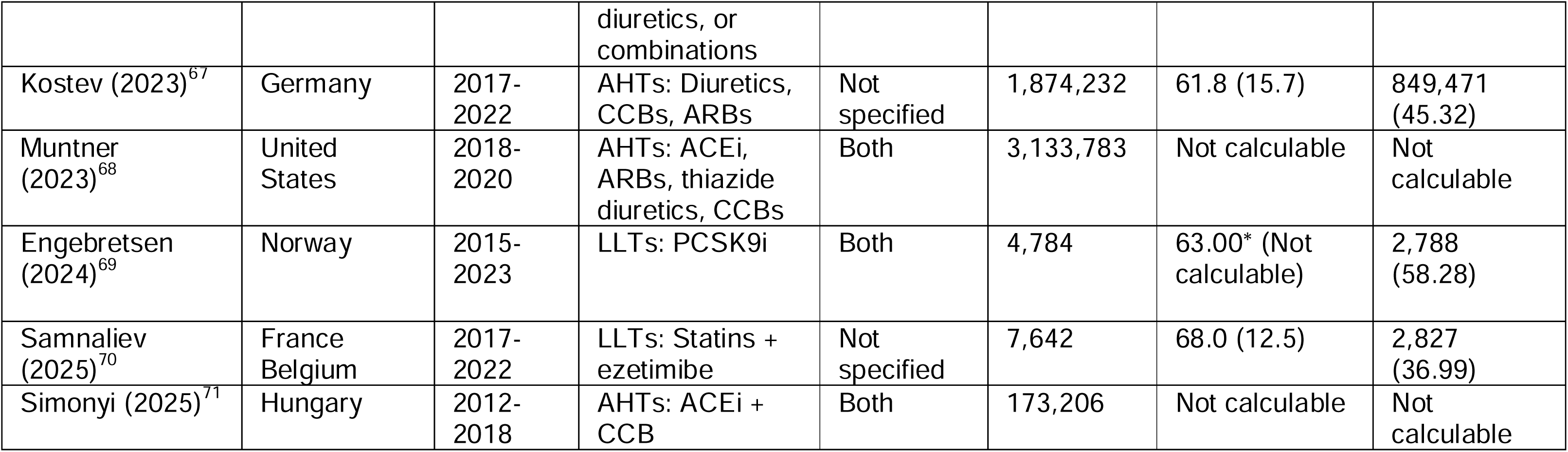
Characteristics of included studies (n = 11,645,049 patient registrations). Simonyi (2015) was a conference abstract but was included as its data matched with results from a non-English journal article. AHTs, antihypertensives; LLTs, lipid-lowering therapies; ARB, angiotensin receptor blockers; ACEi, angiotensin-converting enzyme inhibitors; CCB, calcium channel blockers.

Methodological variations included the specific methods used to compute discontinuation, the length of gap needed to establish a discontinuation ‘event’, and whether medication switching was counted as a discontinuation event (online supplemental file, p9-14).

### Risk of bias assessment

Risk of bias was assessed for all included studies using ROBINS-E.^24^ Most studies had a low risk of bias; twenty two had some concerns about bias (online supplemental file, p15).^29,30,32,33,37,39–41,43,45–49,53,54,57,58,62,66,68,69^ Only two studies had estimates with a very high risk of bias, both due to non-adjustment for age and/or sex.^42,49^ The commonest sources of bias were lack of confounder adjustment or selection bias (e.g. selecting random samples of a larger population without assessing whether its features are similar to the overall study population).

### Outcomes

For both LLTs and AHTs, discontinuation rates varied by cohort demographics, treatment indication (primary versus secondary prevention) and method for ascertaining discontinuation (seven different methods were used, see online supplemental file, p17).

Overall, for LLTs the median (IQR) discontinuation prevalence was 43% (33%–59%).^30,32,41,42,45,46,48–50,52–54,58,59,61–63,66,69^ This was similar for primary (39% [30%–52%]),^32,45,46,49,52,59,61–63^ and secondary prevention (39% [33%–42%]);^42,45,46,48,52,59,61–63^ but higher in mixed prevention (45% [35%–59%]). ^30,41,45,46,50,52–54,58,59,61–63,66,69^ Discontinuation prevalence within a paper increased when medication switches were defined as a discontinuation event, or where shorter gaps in medication supply were permitted (online supplemental file, p9, p18).

Median restarting prevalence after LLT discontinuation was 58% (40%–61%).^32,42,45,52,54,59,66^ and did not appear to vary by primary/ secondary prevention LLT indication (online supplemental file, p15).

For AHTs, overall median discontinuation prevalence was 41% (30%–49%).^33,34,37,43,51,64^ This was similar for exclusively primary prevention indications (43% [41%–47%]);^33,37,64^ but lower for mixed prevention indications (27% [26%–44%]).^34,43,51^ No data was available for secondary prevention.

When including medication switches as discontinuation events, discontinuation prevalence was 49% (26%–60%).^13,33,34,37,44,51,68,71^ Median discontinuation in primary care populations was 64% (57%–64%),^33,37,44^ and 26% (11%–44%) in mixed prevention populations.^13,34,51,68,71^

### Association of age with discontinuation of LLTs and AHTs

Thirteen articles reported LLT discontinuation by age.^30,32,40,42,45,48,49,53,54,58,61,69^ Four that discontinuation was likeliest for the youngest age group (online supplemental file, p19, p25).^40,45,61,69^ However, where the eldest age group was >80 years, discontinuation was highest in this group.^49,54,58^

Two studies with over two age categories found a U-shaped curve in discontinuation for which the youngest and oldest ages were most likely to discontinue,^45,49^ but this trend was not consistent for all studies. In papers with data from primary and secondary prevention, discontinuation trends were generally similar for both treatment indications.^45,61^

Three articles described associations between age and restarting statins. Vinogradova *et al.* (2016) found that UK patients receiving statins for primary prevention were most likely to restart around 55 years old.^45^ For secondary CVD prevention in the UK and US, patients under the age of 55 or 65, respectively, were more likely to restart statins.^42,45^ Ofori-Asenso *et al*. (2019) found that restarting was less likely over the age of 75 years.^58^

For AHTs, five studies reported discontinuation by age.^13,33,37,64,68^ Four reported that the youngest patients were more likely to discontinue than older groups (online supplemental file, p23); ^13,33,64,68^ two of which found a U-shaped association between age and antihypertensive discontinuation similar to statin discontinuation.^13,68^ In contrast, Ah *et al*. (2016) reported that discontinuation risk was highest in patients aged 80 years or over (adjusted hazard ratio (aHR) [95% confidence interval (CI)]; 1.25 [1.17 to 1.34]), relative to patients aged 18 to 64.^37^ Associations between age and discontinuation were consistent across primary and mixed primary/secondary prevention populations, irrespective of how discontinuation was quantified (online supplemental file, 23).

### Association of sex with discontinuation of LLTs and AHTs

11 papers reported sex-specific LLT discontinuation data.^30,40,45,46,48,49,53,54,58,61,69^ Six indicated that discontinuation was more likely in women than men (Figure 2A), including for PCSK9 inhibitor monoclonal antibodies.^45,46,49,53,61,65^ In five other papers, there was no significant association between sex and LLT discontinuation.^30,40,48,53,54,58^Two papers reported sex-specific data on statin restarting: Vinogradova *et al*. (2016) reported that women restarted statin treatment less often than men (primary prevention aHR 0.92 [0.91 to 0.93]; secondary prevention aHR 0.93 [0.91 to 0.95]);^45^ as did Ofori-Asenso *et al.* (2019) (0.96 [0.92 to 0.98]).^58^

**Figure 2.**
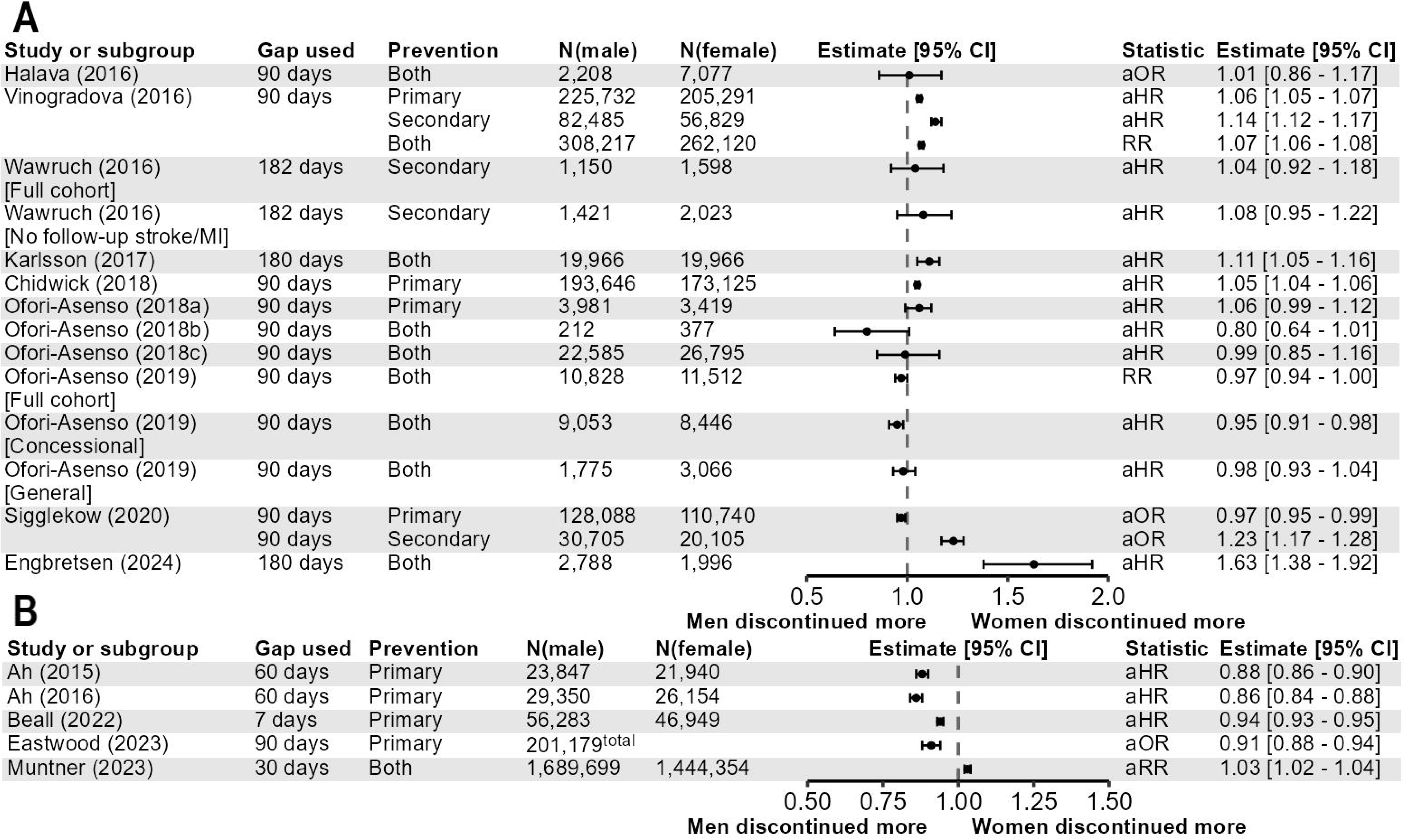
Association of sex with discontinuation of (A) lipid lowering therapies and (B) antihypertensives. Abbreviations: aHR; adjusted hazard ratio; aOR, adjusted odds ratio; RR, risk ratio.

Five articles reported AHT discontinuation by sex (Figure 2B).^13,33,37,64,68^ Four studies in primary prevention populations found that males were more likely to discontinue AHT treatment.^13,33,37,64^ This pattern was reversed in a larger study which included patients using AHTs for both primary and secondary prevention, though the magnitude of the effect estimate was small (adjusted risk ratio [95% CI]; 1.03 [1.02 to 1.04]).^68^

### Association of socioeconomic position with discontinuation of LLTs and AHTs

Four papers reported associations between SEP and LLT discontinuation. Generally, SEP and discontinuation were inversely associated, ^45,46,49,61^ though these associations were of low magnitude in two studies.^45,49^ Associations did not appear to vary by primary versus secondary prevention but were strongest in the study that defined SEP using income.^46^

For AHTs, two papers reported associations between SEP measures and discontinuation (Figure 3B). Beall *et al.* found that patients from neighbourhoods in a higher income quintile were less likely to discontinue an antihypertensive than those in the living in a neighbourhood of the lowest income quintile.^64^ Eastwood *et al.* found no association between practice-level IMD quintile and AHT discontinuation.^13^

**Figure 3.**
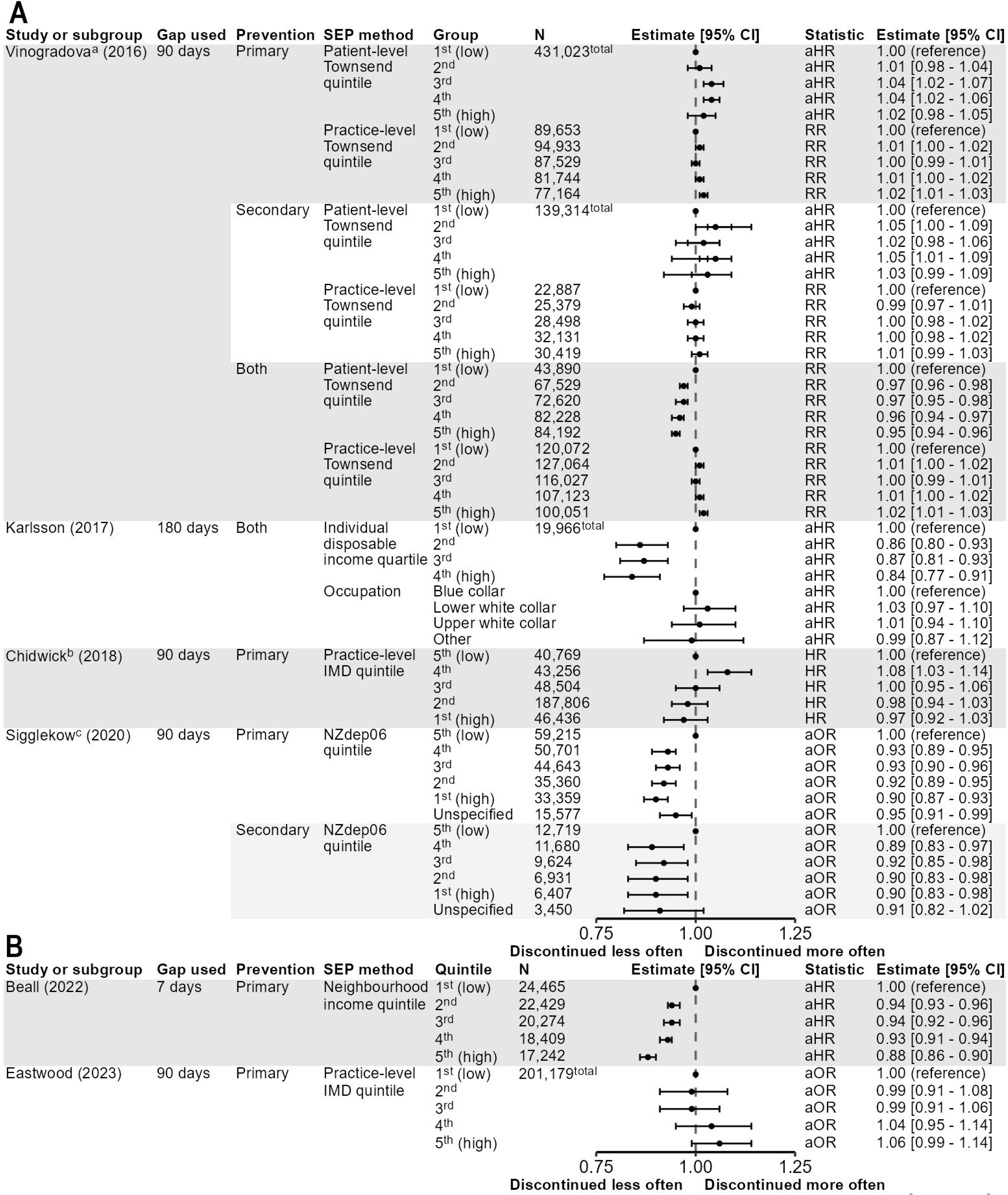
Association of socioeconomic position with discontinuation of (A) lipid-lowering therapies and (B) antihypertensives. ^a,b,c^The plotted estimates/95% CIs for these papers were obtained by changing the reference level from the highest SEP group to the lowest SEP group. Abbreviations: aHR; adjusted hazard ratio; aOR, adjusted odds ratio; RR, risk ratio.

### Association of ethnicity with discontinuation of LLTs and AHTs

This section uses the same terminology to describe ethnic groups as in the referenced papers.

Three papers described associations between LLT discontinuation and ethnicity (Figure 4A).^45,46,61^ In Karlsson *et al*.’s study of type 2 diabetes patients in Sweden, ethnicity was proxied by country of birth. All groups of patients not born in Sweden experienced more discontinuation than patients born in Sweden; this association strengthened with increased geographic distance from Sweden.^46^ Similarly, Vinogradova *et al*. found that all groups of non-White patients were more likely to discontinue statins than their White counterparts, in both primary and secondary CVD prevention.^45^ All non-White ethnic groups in this study were also more likely to restart treatment than White/Not recorded individuals for primary CVD prevention, but only some groups in secondary CVD prevention (data not shown). In Sigglekow *et al*., non-European patients were generally more likely to discontinue for primary and secondary prevention.^61^

**Figure 4.**
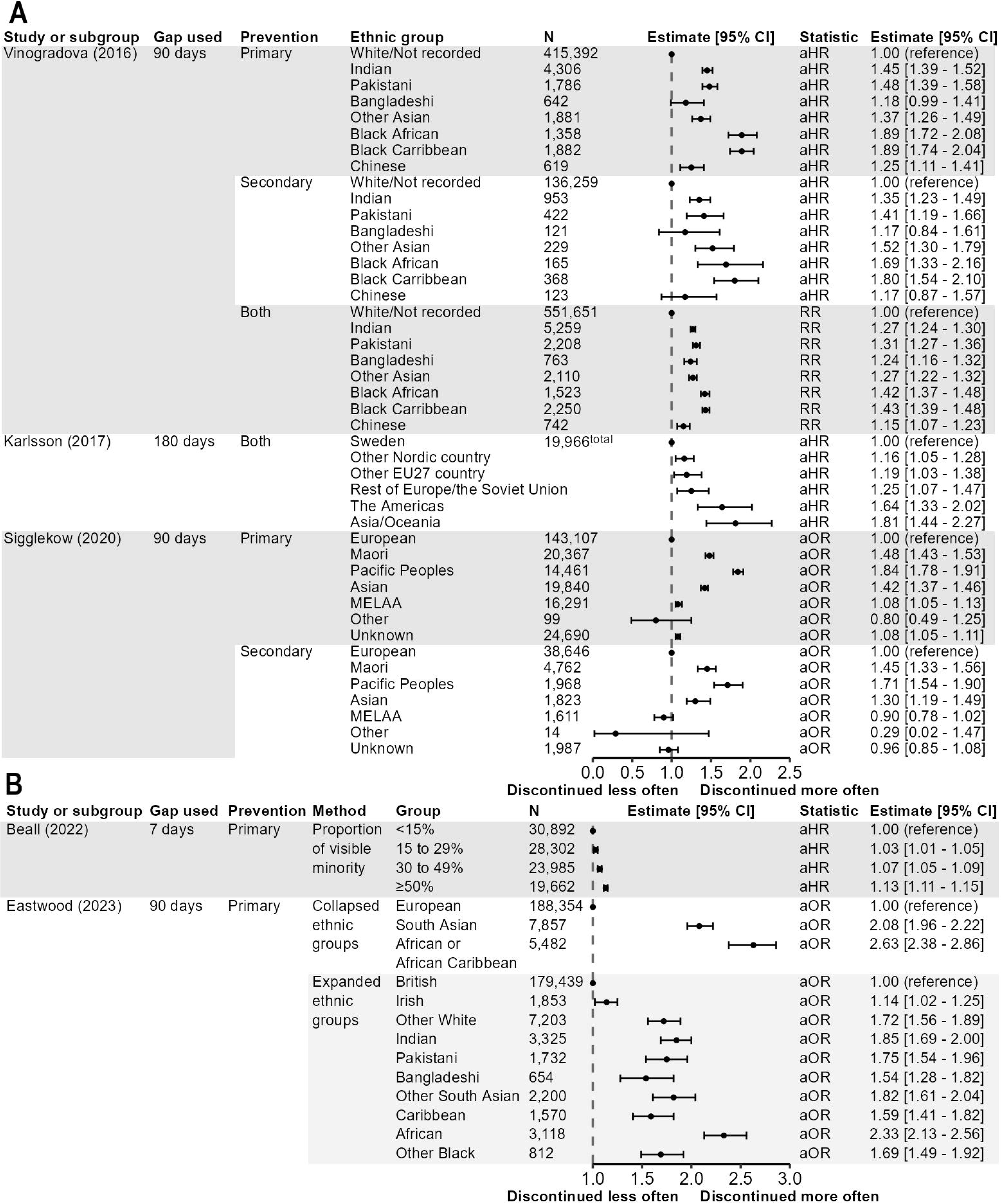
Association of ethnicity with discontinuation of (A) lipid-lowering therapies and (B) antihypertensives. Abbreviations: aHR; adjusted hazard ratio, aOR, adjusted odds ratio.

**Figure 5.** PRISMA flow chart diagram. Abbreviations: aHR; adjusted hazard ratio; aOR, adjusted odds ratio; RR, risk ratio.

Two papers described trends in AHT discontinuation with respect to ethnicity (Figure 4B). Beall *et al.* found that patients in Alberta, Canada, patients using AHTs to treat uncomplicated hypertension were more likely to discontinue as the ‘proportion of visible minority’ (i.e. non-White individuals who are not aboriginal peoples^72^) in their area increased.^64^ In UK primary care data, Eastwood *et al*. found that patients with recorded South Asian (aOR [95% CI] = 2.08 [1.96–2.22]) or African/African Caribbean ethnicity (2.60 [2.38–2.86]) were more likely to discontinue than those with a recorded European ethnicity.^13^ When disaggregated into finer ethnicity definitions, all patients with a code for non-British ethnicity were more likely to discontinue than patients with a code for British ethnicity. Patients with an African ethnicity code had the highest odds of discontinuing (2.33 [2.13–2.56]).

## DISCUSSION

We have found that significant proportions of patients discontinue LLTs or AHTs used in primary care for either primary (up to 64% AHTs; 80% LLTs) or secondary CVD prevention (up to 72% for LLTs). Younger (<60 years) and older patients (>75 years) are more likely to discontinue LLTs or AHTs than patients aged between 60 and 75 years. Restarting statins was more common for secondary CVD prevention patients. Women may be more likely to discontinue LLTs than men for both primary and secondary CVD prevention, but men were more likely to discontinue AHTs for primary CVD prevention. Lower income was associated with discontinuation for both AHTs and LLTs. Discontinuation was more likely in ethnic minority groups.

Previous meta-analyses have found that statin discontinuation was higher for patients with lower income but neither females nor non-White patients.^12,28^ ^9,54^ Discrepancies with our findings may be explained by the use of trial versus real world data. We observed that women were typically more likely to discontinue LLTs than men. This may be partially explained by the higher prevalence of statin-related muscle symptoms and new-onset diabetes in women than in men.^73^ We also observed that men discontinued AHTs more often than women when using them for primary prevention of CVD, but not in a mixed prevention population. Male sexual dysfunction due to AHT use is a common adverse drug event (ADE) and may affect discontinuation, though more research is needed.^74^ In a mixed prevention population, where a proportion of patients have already experienced cardiovascular events, such ADEs may be more tolerable, compared to in primary prevention.

Whilst both AHT and LLT discontinuation were strongly inversely associated with income in two included studies,^46,64^ associations were less clear when multifaceted SEP measures were used.^13,45,49^ The use of area-level SEP measures, which may misclassify individual SEP, may have further diluted associations. Additionally, for systems where most costs are state-subsidised, SEP may not associate as strongly with discontinuation, unlike in privatised systems, where it may impact affordability of and access to care; this could explain differences we found between UK and US data.^13,64^

Belonging to an ethnic minority was associated with increased discontinuation for both AHTs and LLTs,^13,45,46,64^ with absolute risk differences of ≤20%, compared with white groups.^13^ The relationship between ethnicity and discontinuation can easily be confounded by age or SEP, as ethnic minority groups are on average younger and have lower SEP,^13^ factors which are associated with worse persistence. However, the estimates we obtained were adjusted for these factors, suggesting that ethnicity is associated with AHT/LLT discontinuation through other factors, such as experiences of interpersonal racism, or other structural issues which negatively affect care through social and psychological pathways.^75^

Effects may be mediated through systematic factors, such as reduced engagement with healthcare systems or mistrust, as in studies on antidepressant use.^76^ Importantly, the far higher rates of both LLT *and* AHT discontinuation in ethnic minority groups have grave implications for CVD risk in these groups, yet present key modifiable targets.

This is the first review of exclusively EHR-based LLT *and* AHT discontinuation data from real world patients, instead of either RCTs alone or a blend of real world and RCT data. Our data therefore capture patterns in discontinuation and restarting of LLTs and AHTs in primary care which reflect real-world clinical practice. Though methodologically heterogenous, discontinuation patterns for age, and ethnicity were similar across multiple studies, suggesting validity of these patterns. Our simultaneous reporting of LLT and AHT discontinuation supports inference regarding potential drivers, e.g. the sex differences in LLT versus AHT discontinuation suggest sex-specific ADE may be a key causative factor. Further, the similar ethnic patterns in discontinuation for LLT and AHT imply compounded but eminently modifiable disadvantage across many non-European groups. Additionally, whilst not all effects were large, small differences in discontinuation across groups can mediate large shifts in CVD risk at population scales.

Our findings should be considered with respect to this review’s limitations. Causal interpretation of the review data is difficult: both SEP and ethnicity have high degrees of missingness in EHR data, which can bias estimates of the association between these factors and LLT/AHT discontinuation away from the null if SEP or ethnicity are more often recorded in patients with worse illness. EHR-based SEP and ethnicity measures also lack relevant information on engagement with health services or health education. This review also reported on data exclusively from higher-income countries, with no data from lower- or middle-income countries, which limits the generalisability of the results. Our studies were also methodologically heterogeneous, which precluded meta-analysis. Widening or narrowing the allowed ‘gap’ used to define discontinuation can alter estimates by >10% (online supplemental file p9). ^49^

Future researchers should adopt existing discontinuation measures – most likely the ‘grace period’ approach used by most studies included in this review – with a discontinuation gap that allows comparison of discontinuation rates across study populations. Studies which investigate the intersection of SEP and ethnicity would be useful to inform intervention targeting, as would investigation of biological factors in drug discontinuation and qualitative research exploring reasons for discontinuation. The latter is key to explaining differences in medication use across groups.

## CONCLUSIONS

In this contemporary synthesis of real-world data which uniquely combines evidence for LLT and AHT discontinuation, we found a median discontinuation prevalence of 43% (33%–59%) for LLTs and 58% (40%–61%) for AHTs. We report consistent inequalities in discontinuation, including higher discontinuation prevalences for; people at extremes of younger and older age (LLT and AHT), women for LLT and men for AHT, income-based low SEP (LLT and AHT), and minority ethnic groups (LLT and AHT).Future work using large-scale EHR data should use methods consistent with previous work to allow comparability of results. Qualitative work should assess causes for discontinuation, through which interventions for at-risk groups can be designed.

## Data Availability

Study code and summary information from the included papers will be made available by the corresponding author on request.

## DATA AVAILABILITY STATEMENT

Data including pre-harmonised data from papers and the code used to harmonise and visualise data will be made available by the corresponding author on request. Search strategies and the systematic review protocol are included in the Supplemental data.

## ETHICS STATEMENTS

## Patient consent for publication

Not applicable.

## Ethics approval

Not applicable.

## FOOTNOTES

## Contributors

NC, SVE, and AFS conceptualised the study. SP wrote the protocol with input from NC, SVE, and AFS. SP designed the initial search strategy, which was edited according to recommendations from CB. SP performed the literature searches and reviewed the abstracts with NN. SVE adjudicated on disputes regarding study inclusion. SP performed full text screening, risk of bias analyses, data extraction, and data analysis/visualisation. SP wrote the first draft of the manuscript. NC, SVE, AFS contributed to data interpretation. NC, SVE, AFS, CB, and NN contributed to article review. All authors contributed to and have approved of the final article.

## Patient and public involvement

Patients and/or the public were not involved in the design, or conduct, or reporting, or dissemination plans of this research.

