## Supplemental Data for "Discontinuation of antihypertensive and lipid-lowering medication in primary care: a systematic review of observational data and socio-demographic differences"

**Contents**

Page 2: Table S1 – Supplementary methods.

Page 7: Table S2 – PRISMA 2020 checklist.

Page 9: Table S3 – Discontinuation and restarting measures of included studies

Page 15: Table S4 – Risk of bias of included studies.

Page 17: Table S5 – Discontinuation method descriptions and usage by study.

Page 18: Table S6 – Discontinuation and restarting prevalence by study population and medication ‘switching’ as discontinuation.

Page 19: Table S7 – Measures of association between sociodemographic categories and use of ‘switching’ as lipid-lowering therapy discontinuation.

Page 23: Table S8 – Measures of association between sociodemographic categories and antihypertensive discontinuation.

Page 25: Figure S1 – Association of age group with discontinuation of (A) lipid-lowering therapies and (B) antihypertensives.

**Table S1 – Supplementary methods.**

| **Deviations from study protocol** | |
| --- | --- |
| The study protocol (PROSPERO ID: CRD420250599340) details a review of glucose-lowering medications, in addition to lipid-lowering therapies and antihypertensives. We limited our focus in this report to lipid-lowering therapies and antihypertensives, given the range of data available and variables under assessment. The searching and reviewing of abstracts and full-text articles for glucose-lowering medications is currently ongoing, with a similar focus on discontinuation/restarting prevalence and associative measures for discontinuation and either of age, sex, socioeconomic position, or ethnicity. As a result, terms for relevant glucose-lowering drugs are present within the search strategy. | |
| **Search strategy** | |
| EMBASE (via Ovid) | 1. (benazepril or captopril or enalapril or fosinopril or imidapril or lisinopril or perindopril or quinapril or ramipril or trandolapril or azilsartan or candesartan or eprosartan or irbesartan or losartan or olmesartan or telmisartan or valsartan or amlodipine or clevidipine or diltiazem or felodipine or isradipine or lacidipine or lercanidipine or nicardipine or nicardipine o nifedipine or nimodipine or nisoldipine or verapamil or chlortalidone or chlortalidone or chlorthalidone or indapamide or metolazone or bendroflumethiazide or chlorothiazide or chlorthiazide or hydrochlorothiazide or atorvastatin or fluvastatin or lovastatin or pitavastatin or pravastatin or rosuvastatin or simvastatin or ezetimibe or alirocumab or evolocumab or metformin or alogliptin or linagliptin or saxagliptin or sitagliptin or vildagliptin or pioglitazone or glibenclamide or gliclazide or glimepiride or glipizide or glyburide or tolbutamide or bexagliflozin or canagliflozin or dapagliflozin or empagliflozin or ertugliflozin or sotagliflozin or dulaglutide or exenatide or liraglutide or lixisenatide or semaglutide or tirzepatide).ti,ab.  2. (accupril or altace or benazepril or epaned or qbrelis or vasotec or zestril or atacand or avapro or benicar or cozaar or diovan or edarbi or micardis or cardene or cardizem or cartia or katerzia or matzim or norliqva or norvasc or nymalize or procardia or tiadylt or tiazac or verelan or diuril or altoprev or caduet or ezallor or flolipid or lescol or lipitor or livalo or mevacor or nikita or zocor or zypitamag or zetia or praluent or repatha or glumetza or riomet or januvia or nesina or onglyza or tradjenta or zituvio or actos or glucotrol or brenzavvy or farxiga or inpefa or invokana or jardiance or steglatro or bydureon or byetta or mounjaro or ozempic or rybelsus or saxenda or trulicity or victoza or wegovy or zepbound or accuretic or vaseretic or zestoretic or atacand or avalide or azor or benicar or diovan or hyzaar or micardis or edarbyclor or exforge or exforge or tribenzor or azor or caduet or lotrel or aldactazide or lotensin or lypqozet or vytorin or nexlizet or actoplus or invokamet or janumet or jentadueto or kazano or kombiglyze or segluromet or synjardy or trijardy or xigduo or zituvimet or glyxambi or qtern or steglujan or duetact or oseni).ti,ab.  3. (((ACE or angiotensin converting enzyme) adj inhibitor*) or angiotensin receptor blocker* or calcium channel blocker* or thiazide-like diuretic* or thiazide diuretic* or statin* or ((hydroxymethylglutaryl-CoA or HMG-CoA) adj reductase inhibitor*) or ((("proprotein convertase subtilisin/kexin type" adj ("9" or nine)) or PCSK9) adj inhibitor*) or (((dipeptidyl-peptidase adj ("4" or four or IV)) or DPP4) adj inhibitor*) or thiazolidinedione* or sulfonylurea* or sulphonylurea* or (((sodium glucose transporter adj ("2" or two)) or SGLT2) adj inhibitor*) or (((glucagon like peptide adj ("1" or one) adj receptor) or GLP1R) adj agonist*)).ti,ab.  4. (dipeptidyl carboxypeptidase inhibitor/ or angiotensin receptor antagonist/ or calcium channel blocking agent/ or metolazone/ or indapamide/ or thiazide diuretic agent/ or hydroxymethylglutaryl coenzyme A reductase inhibitor/ or PCSK9 inhibitor/ or dipeptidyl peptidase IV inhibitor/ or pioglitazone/ or sulfonylurea derivative/ or sodium glucose cotransporter 2 inhibitor/ or glucagon like peptide 1 receptor agonist/)  5. (((lipid or glucose or blood pressure) adj2 (lowering or reducing)) or anti#hypertensiv* or cholesterol medication$1 or oral hypoglyc$6 or anti#diabeti$4).ti,ab.  6. 1 or 2 or 3 or 4 or 5  7. (((computer?based or computer?stored or electronic or computeri?ed) adj (medical or patient or health) adj (recor* or regist*)) or e$health or ((medical or health) adj information exchange*) or physician order entr$3 or (cohort and (((primary or ambulatory) adj care) or general practice or population based)) or ((health* or prescri* or insur*) adj database)).ti,ab.  8. (CPRD or clinical practice research datalink or THIN or Health Improvement Network or Veteran Affairs or SNDS or "Système National des Données de Santé or Dossier Médical Partagé" or OpenSafely or NHS SDE or national health service secure data environment or Allscripts or athenahealth or Cerner or e-MDs or Epic or EHealth Exchange or CSPI or Meditech or Modernizing Medicine or NextGen or Practice Fusion or Greenway or "Dossier Médical Partagé" or DMP or Centralni zdravstveni informacijski sustav Republike Hrvatske or CEZIH or Zorgi or ZexuzHealth or Chipsoft or CompuGroup Medical or SAP or iSoft or Meierhofer or Nexus or Academisch Huisartsen Ontwikkel Netwerk or AHON or Dutch Data Warehouse).ti,ab.  9. (patient dropouts or contin* or compliance or comply* or discontin* or stopping or restart* or persist* or adher* or concordance).ti,ab.  10. 6 and (7 or 8) and 9  11. randomized controlled trial/  12. (random$ adj3 (allocat$ or assign$ or basis or order$)).ab,ti.  13. ((singl$ or doubl$ or trebl$ or tripl$) adj3 (blind$ or mask$)).ab,ti.  14. observational study/  15. 10 not ((11 or 12 or 13) not 14)  16. limit 15 to english  17. limit 16 to yr="2010-current"  18. limit 17 to human |
| PubMed | 1. benazepril OR captopril OR enalapril OR fosinopril OR imidapril OR lisinopril OR perindopril OR quinapril OR ramipril OR trandolapril OR azilsartan OR candesartan OR eprosartan OR irbesartan OR losartan OR olmesartan OR telmisartan OR valsartan OR amlodipine OR clevidipine OR diltiazem OR felodipine OR isradipine OR lacidipine OR lercanidipine OR nicardipine OR nicardipine OR nifedipine OR nimodipine OR nisoldipine OR verapamil OR chlortalidone OR chlortalidone OR chlorthalidone OR indapamide OR metolazone OR bendroflumethiazide OR chlorothiazide OR chlorthiazide OR hydrochlorothiazide OR atorvastatin OR fluvastatin OR lovastatin OR pitavastatin OR pravastatin OR rosuvastatin OR simvastatin OR ezetimibe OR alirocumab OR evolocumab OR metformin OR alogliptin OR linagliptin OR saxagliptin OR sitagliptin OR vildagliptin OR pioglitazone OR glibenclamide OR gliclazide OR glimepiride OR glipizide OR glyburide OR tolbutamide OR bexagliflozin OR canagliflozin OR dapagliflozin OR empagliflozin OR ertugliflozin OR sotagliflozin OR dulaglutide OR exenatide OR liraglutide OR lixisenatide OR semaglutide OR tirzepatide  2. accupril OR altace OR benazepril OR epaned OR qbrelis OR vasotec OR zestril OR atacand OR avapro OR benicar OR cozaar OR diovan OR edarbi OR micardis OR cardene OR cardizem OR cartia OR katerzia OR matzim OR norliqva OR norvasc OR nymalize OR procardia OR tiadylt OR tiazac OR verelan OR diuril OR altoprev OR caduet OR ezallor OR flolipid OR lescol OR lipitor OR livalo OR mevacor OR nikita OR zocor OR zypitamag OR zetia OR praluent OR repatha OR glumetza OR riomet OR januvia OR nesina OR onglyza OR tradjenta OR zituvio OR actos OR glucotrol OR brenzavvy OR farxiga OR inpefa OR invokana OR jardiance OR steglatro OR bydureon OR byetta OR mounjaro OR ozempic OR rybelsus OR saxenda OR trulicity OR victoza OR wegovy OR zepbound OR accuretic OR vaseretic OR zestoretic OR atacand OR avalide OR azor OR benicar OR diovan OR hyzaar OR micardis OR edarbyclor OR exforge OR exforge OR tribenzor OR azor OR caduet OR lotrel OR aldactazide OR lotensin OR lypqozet OR vytorin OR nexlizet OR actoplus OR invokamet OR janumet OR jentadueto OR kazano OR kombiglyze OR segluromet OR synjardy OR trijardy OR xigduo OR zituvimet OR glyxambi OR qtern OR steglujan OR duetact OR oseni  3. ACE inhibitor OR angiotensin receptor blocker OR calcium channel blocker OR thiazide-like diuretic OR thiazide diuretic OR statin OR HMG-CoA reductase inhibitors OR proprotein convertase subtilisin/kexin type 9 OR PCSK9 inhibitor OR dipeptidyl-peptidase IV inhibitor OR DPP4 inhibitor OR thiazolidinediones OR sulfonylureas OR sulphonylureas OR sodium glucose transporter 2 inhibitor OR SGLT2 inhibitor OR glucagon like peptide 1 receptor agonist OR GLP1R agonist  4. "angiotensin-converting enzyme inhibitors"[MeSH Terms] or "angiotensin receptor antagonists"[MeSH Terms] or "calcium channel blockers"[MeSH Terms] or "metolazone"[MeSH Terms] or "indapamide"[MeSH Terms] or “sodium chloride symporter inhibitors"[MeSH Terms] or "hydroxymethylglutaryl-CoA reductase inhibitors"[MeSH Terms] or "PCSK9 inhibitors"[MeSH Terms] or "dipeptidyl peptidase IV inhibitors"[MeSH Terms] or " thiazolidinediones"[MeSH Terms] or "sulfonylurea compounds"[MeSH Terms] or "sodium glucose transporter 2 inhibitors"[MeSH Terms] or "glucagon-like peptide-1 receptor agonists"[MeSH Terms]  5. "lipid*lowering" OR "glucose*lowering" OR "blood*pressure lowering" OR "lipid*reducing" OR "glucose*reducing" OR "blood*pressure reducing" OR "anti*hypertensiv*" OR "cholesterol medication" OR "oral hypoglyc*mic*" OR "anti*diabeti*"  6. #1 OR #2 OR #3 OR #4 OR #5  7. "electronic health records"[MeSH Terms] OR (("computer*based" OR "computer*stored" OR "computeri*ed" OR "electronic") AND ("medical" OR "patient" OR "health") AND "recor*") OR "order entr*" OR "e-health" OR "ehealth" OR "EHR" OR (("medical" OR "health") AND "information exchange") OR "physician order entr*" OR ("cohort" AND ("primary care" OR "general practice" OR "ambulatory care" OR "population based") OR (("health*" OR "prescri*" OR "insur*") AND "database"))  8. "CPRD" OR "clinical practice research datalink" OR "THIN" OR "Health Improvement Network" OR "Veteran Affairs" OR "SNDS" OR "Système National des Données de Santé" OR "Dossier Médical Partagé" OR "OpenSafely" OR "NHS SDE" OR "national health service secure data environment" OR "Allscripts" OR "athenahealth" OR "Cerner" OR "e-MDs" OR "Epic" OR "EHealth Exchange" OR "CSPI" OR "Meditech" OR "Modernizing Medicine" OR "NextGen" OR "Practice Fusion" OR "Greenway" OR "Dossier Médical Partagé" OR "DMP" OR "Centralni zdravstveni informacijski sustav Republike Hrvatske" OR "CEZIH" OR "Zorgi" OR "ZexuzHealth" OR "Chipsoft" OR "CompuGroup Medical" OR "SAP" OR "iSoft" OR "Meierhofer" OR "Nexus" OR "Academisch Huisartsen Ontwikkel Netwerk" OR "AHON" OR "Dutch Data Warehouse"  9. "Patient dropouts"[MeSH terms] OR "Patient compliance"[MeSH terms] OR "contin*" OR "compliance" OR “comply*" OR "discontin*" OR "stopping" OR "restart*" OR "persist*" OR "adher*" OR "concordance"  10. #6 AND (#7 OR #8) AND #9  11. #10 NOT ("Clinical Trial"[Publication Type] NOT "Observational Study"[Publication Type])  12. #11 AND english[Filter]  13. #12 AND 2010:2024[pdat]  14. #13 AND humans[Filter] |
| Web of Science (All databases) | 1. TS=(benazepril OR captopril OR enalapril OR fosinopril OR imidapril OR lisinopril OR perindopril OR quinapril OR ramipril OR trandolapril OR azilsartan OR candesartan OR eprosartan OR irbesartan OR losartan OR olmesartan OR telmisartan OR valsartan OR amlodipine OR clevidipine OR diltiazem OR felodipine OR isradipine OR lacidipine OR lercanidipine OR nicardipine OR nicardipine OR nifedipine OR nimodipine OR nisoldipine OR verapamil OR chlortalidone OR chlortalidone OR chlorthalidone OR indapamide OR metolazone OR bendroflumethiazide OR chlorothiazide OR chlorthiazide OR hydrochlorothiazide OR atorvastatin OR fluvastatin OR lovastatin OR pitavastatin OR pravastatin OR rosuvastatin OR simvastatin OR ezetimibe OR alirocumab OR evolocumab OR metformin OR alogliptin OR linagliptin OR saxagliptin OR sitagliptin OR vildagliptin OR pioglitazone OR glibenclamide OR gliclazide OR glimepiride OR glipizide OR glyburide OR tolbutamide OR bexagliflozin OR canagliflozin OR dapagliflozin OR empagliflozin OR ertugliflozin OR sotagliflozin OR dulaglutide OR exenatide OR liraglutide OR lixisenatide OR semaglutide OR tirzepatide)  2. TS=(accupril OR altace OR benazepril OR epaned OR qbrelis OR vasotec OR zestril OR atacand OR avapro OR benicar OR cozaar OR diovan OR edarbi OR micardis OR cardene OR cardizem OR cartia OR katerzia OR matzim OR norliqva OR norvasc OR nymalize OR procardia OR tiadylt OR tiazac OR verelan OR diuril OR altoprev OR caduet OR ezallor OR flolipid OR lescol OR lipitor OR livalo OR mevacor OR nikita OR zocor OR zypitamag OR zetia OR praluent OR repatha OR glumetza OR riomet OR januvia OR nesina OR onglyza OR tradjenta OR zituvio OR actos OR glucotrol OR brenzavvy OR farxiga OR inpefa OR invokana OR jardiance OR steglatro OR bydureon OR byetta OR mounjaro OR ozempic OR rybelsus OR saxenda OR trulicity OR victoza OR wegovy OR zepbound OR accuretic OR vaseretic OR zestoretic OR atacand OR avalide OR azor OR benicar OR diovan OR hyzaar OR micardis OR edarbyclor OR exforge OR exforge OR tribenzor OR azor OR caduet OR lotrel OR aldactazide OR lotensin OR lypqozet OR vytorin OR nexlizet OR actoplus OR invokamet OR janumet OR jentadueto OR kazano OR kombiglyze OR segluromet OR synjardy OR trijardy OR xigduo OR zituvimet OR glyxambi OR qtern OR steglujan OR duetact OR oseni)  3. TS=((("ACE" OR "angiotensin converting enzyme") NEAR/0 inhibitor*) OR angiotensin receptor blocker OR calcium channel blocker OR (("thiazide-like" OR "thiazide") NEAR/0 diuretic*) OR statin* OR (("HMG-CoA" OR "hydroxymethylglutaryl-CoA") NEAR/0 reductase inhibitor*) OR (("PCSK9" OR "proprotein convertase subtilisin/kexin type 9") NEAR/0 inhibitor*) OR (("DPP4" OR ("dipeptidyl-peptidase" NEAR/0 ("four" or "4" or "IV"))) NEAR/0 inhibitor*) OR thiazolidinedione* OR sulphonylurea* OR sulfonylurea* OR (("SGLT2" OR "sodium glucose transporter" NEAR/0 ("two" or "2")) NEAR/0 inhibitor*) OR (("GLP1R" OR ("glucagon like peptide" NEAR/1 ("one" or "1") NEAR/0 receptor)) NEAR/0 agonist*))  4. TS=("lipid lowering" OR "glucose lowering" OR "blood pressure lowering" OR "lipid reducing" OR "glucose reducing" OR "blood pressure reducing“ OR "anti$hypertensiv*" OR "cholesterol medication*" OR "oral hypoglyc$emic*" OR "anti$diabet*")  5. #1 OR #2 OR #3 OR #4  6. TS=((("computer based" OR "computer stored" OR "electronic" OR "computeri?ed") NEAR/0 ("medical" OR "patient" OR "health") NEAR/0 ("recor*" OR "regist*")) OR "e$health" OR (("medical" OR "health") NEAR/0 "information exchange") OR "physician order entr*" OR ("cohort" AND ((("primary" or "ambulatory") NEAR/0 care) OR "general practice" OR "population based")) OR (("health*" OR "prescri*" OR "insur*") NEAR/0 "database"))  7. TS=("CPRD" OR "clinical practice research datalink" OR "THIN" or "Health Improvement Network" OR "Veteran Affairs" OR "SNDS" OR "Système National des Données de Santé" OR "Dossier Médical Partagé" OR "DMP" OR "OpenSafely" OR "NHS SDE" OR "national health service secure data environment" OR "Allscripts" OR "athenahealth" OR "Cerner" OR "e-MDs" OR "Epic" OR "EHealth Exchange" OR "CSPI" OR "Meditech" OR "Modernizing Medicine" OR "NextGen" OR "Practice Fusion" OR "Greenway" OR "Centralni zdravstveni informacijski sustav Republike Hrvatske" OR "CEZIH" OR "Zorgi" OR "ZexuzHealth" OR "Chipsoft" OR "CompuGroup Medical" OR "SAP" OR "iSoft" OR "Meierhofer" OR "Nexus" OR "Academisch Huisartsen Ontwikkel Netwerk" OR "AHON" OR "Dutch Data Warehouse")  8. TS=("patient dropouts" OR "contin*" OR "compliance" OR "comply*" OR "discontin*" OR "stopping" OR "restart*" OR "persist*" OR "adher*“ OR "concordance")  9. #5 AND (#6 OR #7) AND #8  10. ALL=("randomi?ed controlled trial*") OR AB=("randomly" OR "placebo" OR (rando* NEAR/1 (allocat* OR assign* OR basis OR order*)) OR ((singl* OR doubl* OR trebl* OR tripl*) NEAR/1 blinded)) OR TI=("randomly" OR "placebo" OR (rando* NEAR/1 (allocat* OR assign* OR basis OR order*)) OR ((singl* OR doubl* OR trebl* OR tripl*) NEAR/1 blinded))  11. #9 NOT (#10 NOT KP=("observational"))  12. #11 AND PY=(2010 OR 2011 OR 2012 OR 2013 OR 2014 OR 2015 OR 2016 OR 2017 OR 2018 OR 2019 OR 2020 OR 2021 OR 2022 OR 2023 OR 2024)  Then restrict #12 to English language only in web interface. |
| CINAHL Plus | S1. benazepril OR captopril OR enalapril OR fosinopril OR imidapril OR lisinopril OR perindopril OR quinapril OR ramipril OR trandolapril OR azilsartan OR candesartan OR eprosartan OR irbesartan OR losartan OR olmesartan OR telmisartan OR valsartan OR amlodipine OR clevidipine OR diltiazem OR felodipine OR isradipine OR lacidipine OR lercanidipine OR nicardipine OR nicardipine OR nifedipine OR nimodipine OR nisoldipine OR verapamil OR chlortalidone OR chlortalidone OR chlorthalidone OR indapamide OR metolazone OR bendroflumethiazide OR chlorothiazide OR chlorthiazide OR hydrochlorothiazide OR atorvastatin OR fluvastatin OR lovastatin OR pitavastatin OR pravastatin OR rosuvastatin OR simvastatin OR ezetimibe OR alirocumab OR evolocumab OR metformin OR alogliptin OR linagliptin OR saxagliptin OR sitagliptin OR vildagliptin OR pioglitazone OR glibenclamide OR gliclazide OR glimepiride OR glipizide OR glyburide OR tolbutamide OR bexagliflozin OR canagliflozin OR dapagliflozin OR empagliflozin OR ertugliflozin OR sotagliflozin OR dulaglutide OR exenatide OR liraglutide OR lixisenatide OR semaglutide OR tirzepatide  S2. accupril OR altace OR benazepril OR epaned OR qbrelis OR vasotec OR zestril OR atacand OR avapro OR benicar OR cozaar OR diovan OR edarbi OR micardis OR cardene OR cardizem OR cartia OR katerzia OR matzim OR norliqva OR norvasc OR nymalize OR procardia OR tiadylt OR tiazac OR verelan OR diuril OR altoprev OR caduet OR ezallor OR flolipid OR lescol OR lipitor OR livalo OR mevacor OR nikita OR zocor OR zypitamag OR zetia OR praluent OR repatha OR glumetza OR riomet OR januvia OR nesina OR onglyza OR tradjenta OR zituvio OR actos OR glucotrol OR brenzavvy OR farxiga OR inpefa OR invokana OR jardiance OR steglatro OR bydureon OR byetta OR mounjaro OR ozempic OR rybelsus OR saxenda OR trulicity OR victoza OR wegovy OR zepbound OR accuretic OR vaseretic OR zestoretic OR atacand OR avalide OR azor OR benicar OR diovan OR hyzaar OR micardis OR edarbyclor OR exforge OR exforge OR tribenzor OR azor OR caduet OR lotrel OR aldactazide OR lotensin OR lypqozet OR vytorin OR nexlizet OR actoplus OR invokamet OR janumet OR jentadueto OR kazano OR kombiglyze OR segluromet OR synjardy OR trijardy OR xigduo OR zituvimet OR glyxambi OR qtern OR steglujan OR duetact OR oseni  S3. (("ACE" OR "angiotensin converting enzyme") N0 inhibitor*) OR angiotensin receptor blocker OR calcium channel blocker OR (("thiazide-like" OR "thiazide") N0 diuretic*) OR statin* OR (("HMG-CoA" OR "hydroxymethylglutaryl-CoA") N0 "reductase inhibitor*") OR (("PCSK9" OR "proprotein convertase subtilisin/kexin type 9") N0 inhibitor*) OR (("DPP4" OR ("dipeptidyl-peptidase" N0 ("4" OR "four" OR "IV"))) N0 inhibitor*) OR thiazolidinedione* OR sulphonylurea* OR sulfonylurea* OR (("SGLT2" OR "sodium glucose transporter" ("2" OR "two")) N0 inhibitor*) OR (("GLP1R" OR ("glucagon like peptide" N0 ("1" OR "one"))) N0 "receptor inhibitor*")  S4. (MH "angiotensin receptor antagonists") OR (MH "calcium channel blockers") OR (MH "metolazone") OR (MH "indapamide") OR (MH "Diuretics, Thiazide") OR (MH " Statins") OR (MH "PCSK9 inhibitors") OR (MH "Dipeptidyl Peptidase 4 Inhibitors") OR (MH "thiazolidinediones") OR (MH "sulfonylurea compounds") OR (MH "sodium-glucose co-transporter 2 inhibitors") OR (MH "glucagon-like peptide-1 receptor agonists")  S5. ((lipid OR glucose OR "blood pressure") N0 (?lowering OR ?reducing)) OR anti#hypertensiv* OR "cholesterol medication*" OR "oral hypoglyc*" OR "anti#diabeti*"  S6. S1 OR S2 OR S3 OR S4 OR S5  S7. (("computer?based" OR "computer?stored" OR "computeri?ed" OR "electronic") N0 ("medical" OR "patient" OR "health") N0 "recor*") OR "e#health" OR "EHR" OR (("medical" OR "health") N0 "information exchange") OR "physician order entr*" OR ("cohort" AND ("primary care" OR "general practice" OR "ambulatory care" OR "population based")) OR (("health*" OR "prescri*" OR "insur*") N0 "database"))  S8. "CPRD" OR "clinical practice research datalink" OR "THIN" OR "Health Improvement Network" OR "Veteran Affairs" OR "SNDS" OR "Système National des Données de Santé" OR "Dossier Médical Partagé" OR "OpenSafely" OR "NHS SDE" OR "national health service secure data environment" OR "Allscripts" OR "athenahealth" OR "Cerner" OR "e-MDs" OR "Epic" OR "EHealth Exchange" OR "CSPI" OR "Meditech" OR "Modernizing Medicine" OR "NextGen" OR "Practice Fusion" OR "Greenway" OR "Dossier Médical Partagé" OR "DMP" OR "Centralni zdravstveni informacijski sustav Republike Hrvatske" OR "CEZIH" OR "Zorgi" OR "ZexuzHealth" OR "Chipsoft" OR "CompuGroup Medical" OR "SAP" OR "iSoft" OR "Meierhofer" OR "Nexus" OR "Academisch Huisartsen Ontwikkel Netwerk" OR "AHON" OR "Dutch Data Warehouse"  S9. (MH "Patient Dropouts") OR "contin*" OR "compliance" OR "comply*" OR "discontin*" OR "stopping" OR "restart*" OR "persist*" OR "adher*" OR "concordance"  S10. S6 AND (S7 OR S8) AND S9  S11. (MH "Randomized Controlled Trials+")  S12. TI (random$ N2 (allocate* OR assign* OR basis OR order*)) OR (AB (random$ N2 (allocate* OR assign* OR basis OR order*))  S13. S10 not ((S11 or S12) not (MH "nonexperimental studies"))  S14. S13 **(with limiters: Publication year 2010-)**  S15. S14  **(with limiters: Publication year 2010-; Human)**  S16. S15  **(with limiters: Publication year 2010-; Human; Language: English)** |
| **Search timing** | |
| Initial searches were carried out on the 10^th^ of December 2024; later searches were carried out on the 7^th^ of August 2025. | |
| **Drug classes** | |
| Statins, ezetimibe, and proprotein convertase subtilisin/kextin type 9 inhibitor monoclonal antibodies for lipid-lowering therapies^1–4^, and angiotensin converting enzyme inhibitors, angiotensin 2 receptor blockers, calcium channel blockers, and thiazide/thiazide-like diuretics for antihypertensives.^5–7^ | |
| **Multiple drug classes** | |
| Articles which reported discontinuation data separately for multiple drug classes were included if at least one result related to one of the drug classes listed above. Articles were excluded if they reported combined discontinuation rates including data from drug classes not listed above. | |
| **Data harmonisation – study characteristics** | |
| Age data were extracted as provided if given as a whole-cohort mean and standard deviation. If a paper gave a median and IQR, we used these data to estimate a mean and standard deviation.^8^ If a paper gave a means and standard deviation for multiple subcohorts, we collapsed these into an overall cohort mean and standard deviation using the formulae from the Cochrane handbook.^9^ | |
| **Data harmonisation – reference levels of categorical variables** | |
| For visualisations, we wanted to set the reference level for categorical variables to be similar across all studies. For variables with two levels, such as binary sex variables, this requires taking the inverse of the point estimate and 95% confidence interval (CI) to switch the reference level.  For variables with >2 levels, for example socioeconomic position, we recomputed the 95% confidence intervals and point estimates based on reported values, without using the original data from the paper to fully re-run the modelling. We did this by dividing the point estimate and 95% CI values for the existing non-reference categories by the point estimate of the new reference category. | |

**Table S2 – PRISMA 2020 checklist.**

| **Section and Topic** | **Item #** | **Checklist item** | **Location where item is reported** |
| --- | --- | --- | --- |
| **TITLE** | | |  |
| Title | 1 | Identify the report as a systematic review. | 1 |
| **ABSTRACT** | | |  |
| Abstract | 2 | See the PRISMA 2020 for Abstracts checklist. | 2 |
| **INTRODUCTION** | | |  |
| Rationale | 3 | Describe the rationale for the review in the context of existing knowledge. | 3 |
| Objectives | 4 | Provide an explicit statement of the objective(s) or question(s) the review addresses. | 3 |
| **METHODS** | | |  |
| Eligibility criteria | 5 | Specify the inclusion and exclusion criteria for the review and how studies were grouped for the syntheses. | 4 |
| Information sources | 6 | Specify all databases, registers, websites, organisations, reference lists and other sources searched or consulted to identify studies. Specify the date when each source was last searched or consulted. | 4 |
| Search strategy | 7 | Present the full search strategies for all databases, registers and websites, including any filters and limits used. | S1 Table |
| Selection process | 8 | Specify the methods used to decide whether a study met the inclusion criteria of the review, including how many reviewers screened each record and each report retrieved, whether they worked independently, and if applicable, details of automation tools used in the process. | 4 |
| Data collection process | 9 | Specify the methods used to collect data from reports, including how many reviewers collected data from each report, whether they worked independently, any processes for obtaining or confirming data from study investigators, and if applicable, details of automation tools used in the process. | 5 |
| Data items | 10a | List and define all outcomes for which data were sought. Specify whether all results that were compatible with each outcome domain in each study were sought (e.g. for all measures, time points, analyses), and if not, the methods used to decide which results to collect. | 5 |
|  | 10b | List and define all other variables for which data were sought (e.g. participant and intervention characteristics, funding sources). Describe any assumptions made about any missing or unclear information. | 5 |
| Study risk of bias assessment | 11 | Specify the methods used to assess risk of bias in the included studies, including details of the tool(s) used, how many reviewers assessed each study and whether they worked independently, and if applicable, details of automation tools used in the process. | 5 |
| Effect measures | 12 | Specify for each outcome the effect measure(s) (e.g. risk ratio, mean difference) used in the synthesis or presentation of results. | Tables S6, S7, Figures 2-5, S1 |
| Synthesis methods | 13a | Describe the processes used to decide which studies were eligible for each synthesis (e.g. tabulating the study intervention characteristics and comparing against the planned groups for each synthesis (item #5)). | NA |
|  | 13b | Describe any methods required to prepare the data for presentation or synthesis, such as handling of missing summary statistics, or data conversions. | NA |
|  | 13c | Describe any methods used to tabulate or visually display results of individual studies and syntheses. | NA |
|  | 13d | Describe any methods used to synthesize results and provide a rationale for the choice(s). If meta-analysis was performed, describe the model(s), method(s) to identify the presence and extent of statistical heterogeneity, and software package(s) used. | NA |
|  | 13e | Describe any methods used to explore possible causes of heterogeneity among study results (e.g. subgroup analysis, meta-regression). | NA |
|  | 13f | Describe any sensitivity analyses conducted to assess robustness of the synthesized results. | NA |
| Reporting bias assessment | 14 | Describe any methods used to assess risk of bias due to missing results in a synthesis (arising from reporting biases). | NA |
| Certainty assessment | 15 | Describe any methods used to assess certainty (or confidence) in the body of evidence for an outcome. | NA |
| **RESULTS** | | |  |
| Study selection | 16a | Describe the results of the search and selection process, from the number of records identified in the search to the number of studies included in the review, ideally using a flow diagram. | Figure 1 |
|  | 16b | Cite studies that might appear to meet the inclusion criteria, but which were excluded, and explain why they were excluded. | 7 |
| Study characteristics | 17 | Cite each included study and present its characteristics. | 7-8 |
| Risk of bias in studies | 18 | Present assessments of risk of bias for each included study. | 9-10 |
| Results of individual studies | 19 | For all outcomes, present, for each study: (a) summary statistics for each group (where appropriate) and (b) an effect estimate and its precision (e.g. confidence/credible interval), ideally using structured tables or plots. | 11-16, Figures 2-4, S1, Tables S6, S7 |
| Results of syntheses | 20a | For each synthesis, briefly summarise the characteristics and risk of bias among contributing studies. | 11-16 |
|  | 20b | Present results of all statistical syntheses conducted. If meta-analysis was done, present for each the summary estimate and its precision (e.g. confidence/credible interval) and measures of statistical heterogeneity. If comparing groups, describe the direction of the effect. | NA |
|  | 20c | Present results of all investigations of possible causes of heterogeneity among study results. | NA |
|  | 20d | Present results of all sensitivity analyses conducted to assess the robustness of the synthesized results. | NA |
| Reporting biases | 21 | Present assessments of risk of bias due to missing results (arising from reporting biases) for each synthesis assessed. | NA |
| Certainty of evidence | 22 | Present assessments of certainty (or confidence) in the body of evidence for each outcome assessed. | NA |
| **DISCUSSION** | | |  |
| Discussion | 23a | Provide a general interpretation of the results in the context of other evidence. | 13-14 |
|  | 23b | Discuss any limitations of the evidence included in the review. | 14-15 |
|  | 23c | Discuss any limitations of the review processes used. | 14 |
|  | 23d | Discuss implications of the results for practice, policy, and future research. | 15 |
| **OTHER INFORMATION** | | |  |
| Registration and protocol | 24a | Provide registration information for the review, including register name and registration number, or state that the review was not registered. | 4 |
|  | 24b | Indicate where the review protocol can be accessed, or state that a protocol was not prepared. | 4 |
|  | 24c | Describe and explain any amendments to information provided at registration or in the protocol. | 6 |
| Support | 25 | Describe sources of financial or non-financial support for the review, and the role of the funders or sponsors in the review. | 5 |
| Competing interests | 26 | Declare any competing interests of review authors. | 5 |
| Availability of data, code and other materials | 27 | Report which of the following are publicly available and where they can be found: template data collection forms; data extracted from included studies; data used for all analyses; analytic code; any other materials used in the review. | Supplemental data and data availability statement |

**Table S3 – Discontinuation and restarting measures of included studies.**

| **Title** | **First author (Year)** | **PMID** | **Method** | **Prevention type** | **Gap used (days)** | **Switch as discont.** | **Number initiating** | **Number discontinued** | **Percentage discontinued (2 d.p.)** | **Number restarted** | **Percentage restarted (2 d.p.)** | **Notes** |
| --- | --- | --- | --- | --- | --- | --- | --- | --- | --- | --- | --- | --- |
| Persistence with Antihypertensive Medications in Uncomplicated Treatment-Naive Patients: Effects of Initial Therapeutic Classes | Ah (2015)^10^ | 26713055 | Grace period | Primary | 60 | Yes | 40,692 | 26,063 | 64.05 | Not calculable | Not calculable | Computed total number of participants, mean/SD of age, and total/proportion of males. |
|  |  |  |  |  |  | No |  | 17,384 | 42.72 | Not calculable | Not calculable |  |
| Change in prescription pattern as a potential marker for adverse drug reactions of angiotensin converting enzyme inhibitors | Mahmoudpour (2015)^11^ | 26159317 | Grace period | Both | 91 | No | 1,132 | 308 | 27.2 | Not calculable | Not calculable | Computed total number of males. |
|  |  |  |  |  |  | Yes |  | 629 | 55.6 | Not calculable | Not calculable |  |
| Persistence of fixed and free combination of ramipril and amlodipine in hypertension | Simonyi (2015)^12,13^ | N/A | Grace period | Not specified | Not specified | Not specified | 30,545 | 14,405 | 47.16 | Not calculable | Not calculable | Conference abstract with corresponding Hungarian article (doi: 10.1556/oh.2014.30037). |
| Influence of initial angiotensin receptor blockers on treatment persistence in uncomplicated hypertension: A nation-wide population-based study | Ah (2016)^14^ | 27028796 | Grace period | Primary | 60 | No | 55,504 | 22,137 | 39.88 | Not calculable | Not calculable | Data shown here are for discontinuation of the index ARB used. |
|  |  |  |  |  |  | Yes |  | 27,415 | 49.39 | Not calculable | Not calculable |  |
| Longitudinal treatment patterns among US patients with atherosclerotic cardiovascular disease or familial hypercholesterolemia initiating lipid-lowering pharmacotherapy | Burke (2016)^15^ | 27919365 | Grace period | Both | 90 | No | 92,621 | 39,086 | 42.2 | Not calculable | Not calculable |  |
|  |  |  |  |  |  | Yes |  | 48,626 | 52.5 | Not calculable | Not calculable |  |
| Patterns of statin use and cholesterol goal attainment in a high-risk cardiovascular population: A retrospective study of primary care electronic medical records | García-Gil (2016)^16^ | 26892130 | Post-follow-up window | Primary | 183 | Yes | 21,636 | 6,478 | 29.94 | Not calculable | Not calculable | Computed number of males. Study authors performed imputation for "total cholesterol, LDL-C, triglycerides, glucose, systolic blood pressure, diastolic blood pressure, and BMI". |
| Predictors of first-year statin medication discontinuation: A cohort study | Halava (2016)^17^ | 27578131 | Single prescription only | Both | Not applicable | Not applicable | 9,285 | 1,142 | 12.30 | Not calculable | Not calculable |  |
| Patterns of statin use in a real-world population of patients at high cardiovascular risk | Lin (2016)^18^ | 27231796 | Grace period | Both | 90 | No | 463,707 | 245,765 | 53.0 | Not calculable | Not calculable | Computed mean/SD of age, number/proportion of males, and number of discontinuers. |
| A US Claims-Based Analysis of Real-World Lipid-Lowering Treatment Patterns in Patients With High Cardiovascular Disease Risk or a Previous Coronary Event | Quek (2016)^19^ | 26742468 | Grace period | Secondary | 60 | No | 212,278 | 83,454 | 39.31 | 50,974 | 61.08 | Computed mean/SD of age and numbers/proportions of males, discontinuers, and restarters. |
|  |  |  |  |  |  | Yes |  | 118,553 | 55.99 |  | 42.89 |  |
| How does prescribing for antihypertensive products stack up against guideline recommendations? An Australian population-based study (2006–2014) | Schaffer (2016)^20^ | 27302475 | Grace period | Both | 60 | No | 33,657 | 20,264 | 60.21 | Not calculable | Not calculable | The sample reported here excludes users of beta-blockers, which are included in the full study sample. Computed total counts/proportions of males and discontinuers. |
| Ramipril + amlodipine and ramipril + hydrochlorothiazide fixed-dose combinations in relation to patient adherence | Simonyi (2016)^21^ | 27435392 | Grace period | Primary | 60 | Yes | 39,095 | 25,169 | 64.38 | Not calculable | Not calculable | Discontinuation in the ACEi+DIU group higher than in the ACEi+CCB group. |
| Discontinuation and restarting in patients on statin treatment: prospective open cohort study using a primary care database | Vinogradova (2016)^22^ | 27353261 | Grace period | Both | 90 | No | 570,337 | 262,413 | 46.01 | 190,516 | 72.6 | Computed mean/SD of age and numbers/proportions of males, discontinuers, and restarters for 'Both' row. |
|  |  |  |  | Primary |  |  | 431,023 | 204,662 | 47.48 | 147,305 | 71.97 |  |
|  |  |  |  | Secondary |  |  | 139,314 | 57,791 | 41.48 | 43,211 | 74.77 |  |
| Refill adherence and persistence to lipid-lowering medicines in patients with type 2 diabetes: A nation-wide register-based study | Karlsson (2017)^23^ | 28799214 | Maximum gap | Both | 180 | No | 97,595 | 37,931 | 38.87 | Not calculable | Not calculable | Computed proportion of discontinuing patients. Presented data are after two years' follow-up. |
|  |  |  |  | Primary |  |  | 75,464 | 29,298 | 38.82 | Not calculable | Not calculable |  |
|  |  |  |  | Secondary |  |  | 22,131 | 8,633 | 39.01 | Not calculable | Not calculable |  |
| Medication compliance and clinical outcomes of fixed-dose combinations vs free combinations of an angiotensin II receptor blocker and a calcium channel blocker in hypertension treatment | Tung (2017)^24^ | 28560786 | Not specified | Primary | Not specified | Not specified | 5,680 | Not calculable | Not calculable | Not calculable | Not calculable | Discontinuation in this study was reported as the median number of days before discontinuation. No discontinuation number or proportion was reported. |
| Patient-related characteristics associated with non-persistence with statin therapy in elderly patients following an ischemic stroke | Wawruch (2017)^25^ | 27935151 | Grace period | Secondary | 182 | No | 2,748 | 1092 | 39.73 | Not calculable | Not calculable | Data presented here are for the sub-cohort with no stroke/MI during the follow-up. No patients with stroke/MI during follow-up discontinued. |
| Statin use in cancer survivors versus the general population: Cohort study using primary care data from the UK clinical practice research datalink | Chidwick (2018)^26^ | 30348123 | Grace period | Primary | 90 | No | 366,771 | 189,806 | 51.75 | Not calculable | Not calculable | Computed mean/SD of age and numbers/proportions of males and discontinuers. |
| Utilization Patterns of Lipid-lowering Therapies in Patients With Atherosclerotic Cardiovascular Disease or Diabetes: A Population-based Study in South Korea | Kim (2018)^27^ | 29735297 | Grace period | Both | 90 | No | 1,399,872 | 497,604 | 35.55 | Not calculable | Not calculable | Computed mean/SD of age, number/proportion of males, and number/proportion of discontinuers. |
| Prescription patterns of angiotensin‐converting enzyme inhibitors for various indications: A UK population‐based study | Mahmoudpour (2018)^28^ | 29943849 | Grace period | Both | 183 | No | 254,002 | 63,369 | 24.95 | 17,693 | 27.92 |  |
|  |  |  |  |  |  | Yes |  | 100,790 | 39.68 |  | 17.55 |  |
| Persistence with statins in primary prevention of cardiovascular disease: findings from a cohort of Spanish workers | Malo (2018)^29^ | 28473266 | Relative window | Primary | 2x length of previous prescription | No | 725 | 214 | 29.50 | 124 | 57.90 | Computed number of discontinuers and number of restarters. |
| Treatment Patterns, Statin Intolerance, and Subsequent Cardiovascular Events Among Japanese Patients With High Cardiovascular Risk Initiating Statin Therapy | Nagar (2018)^30^ | 29276211 | Grace period | Both | 60 | No | 15,552 | 5,075 | 32.63 | 736 | 14.50 | Computed mean/SD of age, and numbers/proportions of males, discontinuers, and restarters. |
|  |  |  |  |  |  | Yes |  | 6,222 | 40.01 |  | 11.83 |  |
|  |  |  |  | Primary | 60 | No | 10,250 | 3,331 | 32.50 | 492 | 14.77 |  |
|  |  |  |  |  |  | Yes |  | 4,069 | 39.70 |  | 12.09 |  |
|  |  |  |  | Secondary | 60 | No | 5,302 | 1,744 | 32.89 | 244 | 13.99 |  |
|  |  |  |  |  |  | Yes |  | 2,153 | 40.61 |  | 11.33 |  |
| Patterns of statin use and long-term adherence and persistence among older adults with diabetes | Ofori-Asenso (2018)^31^ | 29658177 | Grace period | Both | 45 | No | 7,400 | 5,873 | 79.36 | Not calculable | Not calculable | Computed number/proportion of males. |
|  |  |  |  |  | 90 |  |  | 4,942 | 66.78 | Not calculable | Not calculable |  |
|  |  |  |  |  | 180 |  |  | 4,044 | 54.65 | Not calculable | Not calculable |  |
| Prevalence and Incidence of Statin Use and 3-Year Adherence and Discontinuation Rates Among Older Adults With Dementia | Ofori-Asenso (2018)^32^ | 29991271 | Grace period | Both | 45 | No | 589 | 279 | 47.37 | Not calculable | Not calculable | Presented discontinuation results are at 3 years' follow-up. Computed number/proportion of males. |
|  |  |  |  |  | 90 |  |  | 346 | 58.74 | Not calculable | Not calculable |  |
|  |  |  |  |  | 180 |  |  | 418 | 70.97 | Not calculable | Not calculable |  |
| Switching, Discontinuation, and Reinitiation of Statins Among Older Adults | Ofori-Asenso (2018)^33^ | 30466527 | Grace period | Both | 90 | No | 49,380 | 31,407 | 63.60 | 18,977 | 60.40 | Computed number/proportion of males. |
|  |  |  |  |  |  | Yes |  | 41,227 | 83.49 | Not calculable | Not calculable |  |
| Fixed-dose combination antihypertensive medications, adherence, and clinical outcomes: A population-based retrospective cohort study | Verma (2018)^34^ | 29889841 | Relative window | Not specified | 1.5x median prescription length | Yes | 13,350 | 11,465 | 85.88 | 3,108 | 27.11 | Computed mean/SD of age and totals/proportions of males, discontinuers, and restarters. |
| Treatment patterns in hyperlipidaemia patients based on administrative claim databases in Japan | Wake (2018)^35^ | 29604481 | Relative window | Not specified | 60 | No | 65,407 | 33,524 | 51.25 | Not calculable | Not calculable | Computed mean/SD of age, and number initiating/discontinuing. |
|  |  |  |  |  |  | Yes |  | 35,331 | 54.02 | Not calculable | Not calculable |  |
| Non-adherence to cardiovascular drugs in older patients with depression: A population-based cohort study | Holvast (2019)^36^ | 30249352 | Grace period | Not specified | 182 | Yes | 1,330 | 555 | 41.73 | Not calculable | Not calculable | Computed total number of participants, mean/SD of age, and totals/proportions of males and discontinuers. Total/matched populations different in size to extent that age/sex values probably inaccurate for initiators. |
| Adherence to and Persistence With Statin Therapy in a Veteran Population | Morotti (2019)^37^ | 30084646 | Grace period | Not specified | 135 | Yes | 164,687 | 56,652 | 34.4 | Not calculable | Not calculable | Data presented here are from 405 days' follow-up. Computed proportion of males and discontinuing patients. |
| Predictors of first‐year nonadherence and discontinuation of statins among older adults: a retrospective cohort study | Ofori-Asenso (2019)^38^ | 30402916 | Grace period | Both | 90 | No | 22,340 | 9,982 | 44.68 | Not calculable | Not calculable | Computed number/proportion of males and number/proportion of discontinuers. |
| Treatment patterns of lipid-lowering therapies and possible statin intolerance among statin users with clinical atherosclerotic cardiovascular disease (ASCVD) or diabetes mellitus (DM) in Taiwan | Chen (2020)^39^ | 31646715 | Grace period | Both | 60 | No | 42,192 | 23,859 | 59.64 | 5,674 | 23.78 | Full study looked at more patients, but only reported demographic information for a subset of ASCVD and DM patients. Computed mean/SD of age, and numbers/proportions of males, discontinuers, and restarters. |
|  |  |  |  |  |  | Yes |  | 35,586 | 84.34 |  | 15.94 |  |
|  |  |  |  | Primary |  | No | 31,100 | 17,869 | 57.46 | 4421 | 24.74 |  |
|  |  |  |  |  |  | Yes |  | 26,745 | 86.00 |  | 16.53 |  |
|  |  |  |  | Secondary |  | No | 11,092 | 5,990 | 54.00 | 1253 | 20.92 |  |
|  |  |  |  |  |  | Yes |  | 8,841 | 79.71 |  | 14.17 |  |
| Real-world evaluation of the impact of statin intensity on adherence and persistence to therapy: A Scottish population-based study | Rezende Macedo do Nascimento (2020)^40^ | 32353163 | Grace period | Both | 60 | Yes | 73,716 | 46,370 | 62.90 | 33,543 | 72.34 | For the grace period result, the data at one year are reported here; discontinuation rates were also recorded at 6 months, 2 years, and 3 years. Computed number discontinuing at one year for the anniversary method. |
|  |  |  |  | Primary |  |  | 63,084 | 41,264 | 65.41 | 29,680 | 71.93 |  |
|  |  |  |  | Secondary |  |  | 10,632 | 5,106 | 48.02 | 3,863 | 75.66 |  |
|  |  |  | Anniversary window | Both | 60* | Yes | 73,716 | 51,527 | 69.90 | Not calculable | Not calculable |  |
| Statin adherence is lower in primary than secondary prevention: A national follow-up study of new users | Sigglekow (2020)^41^ | 33211724 | Grace period | Both | 90 | No | 289,666 | 81,189 | 28.03 | Not calculable | Not calculable | Computed mean/SD of age and number/proportion of males. |
|  |  |  |  | Primary |  |  | 238,855 | 71,179 | 29.80 | Not calculable | Not calculable |  |
|  |  |  |  | Secondary |  |  | 50,811 | 10,010 | 19.70 | Not calculable | Not calculable |  |
| One-year statin persistence and adherence in adults with HIV in the United States | Crockett (2021)^42^ | 33341376 | Anniversary window | Both | 60 | Yes | 28,095 | 8,068 | 28.72 | Not calculable | Not calculable | Computed numbers/proportions of males and discontinuers. |
| Comparison of adherence, persistence, and clinical outcome of generic and brand-name statin users: A retrospective cohort study using the Japanese claims database | Gao (2021)^43^ | 33371973 | Grace period | Both | 90 | No | 28,626 | 7,435 | 25.97 | Not calculable | Not calculable | Computed mean/SD of age, and numbers/proportions of males and discontinuers. |
|  |  |  |  | Primary |  |  | 24,777 | 6,883 | 27.78 | Not calculable | Not calculable |  |
|  |  |  |  | Secondary |  |  | 3,849 | 552 | 14.34 | Not calculable | Not calculable |  |
| Impact of changing reimbursement criteria on statin treatment patterns among patients with atherosclerotic cardiovascular disease or cardiovascular risk factors | Hsu (2021)^44^ | 33180353 | Grace period | Both | 60 | No | 650,815 | 412,179 | 63.33 | Not calculable | Not calculable | Data presented here are from only the more contemporary portion of the study, which occurred from 2014-2016. Computed mean/SD of age, proportion of males, and number initiating/discontinuing. |
|  |  |  |  |  |  | Yes |  | 492,332 | 75.65 | Not calculable | Not calculable |  |
|  |  |  |  | Primary |  | No | 610,411 | 389,270 | 63.77 | Not calculable | Not calculable |  |
|  |  |  |  |  |  | Yes |  | 463,362 | 75.91 | Not calculable | Not calculable |  |
|  |  |  |  | Secondary |  | No | 40,404 | 22,909 | 56.70 | Not calculable | Not calculable |  |
|  |  |  |  |  |  | Yes |  | 28,970 | 71.70 | Not calculable | Not calculable |  |
| Laboratory testing and antihypertensive medication adherence following initial treatment of incident, uncomplicated hypertension: A real-world data analysis | Beall (2022)^45^ | 36125169 | Grace period | Primary | 7 | No | 73,373 | 37,891 | 51.64 | Not calculable | Not calculable | Computed total numbers initiating and total/proportion who discontinued. The number of people initiating vs. the number of total patient population is very different, so the age/sex estimates for the intiators may differ. |
| Gaps and discontinuation of statin treatment in Norway: potential for optimizing management of lipid lowering drugs | Engebretsen (2022)^46^ | 36440353 | Grace period | Not specified | 30, 90, 180, 365, 545, 730 | Yes | 831,097 | 257,640 | 31.00 | Not calculable | Not calculable | Presented discontinuation result is for 90 days' follow-up. |
| Switching, Persistence and Adherence to Statin Therapy: a Retrospective Cohort Study Using the Australian National Pharmacy Data | Talic (2022)^47^ | 34097194 | Grace period | Both | 90 | No | 141,062 | 82,944 | 58.80 | 45,785 | 55.20 | Computed number discontinuing and restarting. |
| Ethnic differences in hypertension management, medication use and blood pressure control in UK primary care, 2006–2019: a retrospective cohort study | Eastwood (2023)^48^ | 36818236 | Maximum gap | Both | 90 | Yes | 201,179 | 19,113 | 9.50 | Not calculable | Not calculable | Computed mean/SD of age, and overall number/proportion of males. |
| Persistence with first-line antihypertensive therapy in Germany: A retrospective cohort study with 2,801,469 patients | Kostev (2023)^49^ | 36871244 | Grace period | Not specified | 180 | Yes | 1,874,232 | 1,337,330 | 71.35 | Not calculable | Not calculable | Data presented here are for 1 year of follow-up; the paper also had data at two and three years of follow-up. Computed mean/SD of age, proportion of males, and overall number/proportion discontinuing. |
| Discontinuation of Renin–Angiotensin System Inhibitors During the Early Stage of the COVID-19 Pandemic | Muntner (2023)^50^ | 36960855 | Grace period | Both | 30 | Yes | 3,133,783 | 374,040 | 11.94 | Not calculable | Not calculable | Participants were divided across six subcohorts, but could appear in each subcohort. Therefore the 'whole' cohort may include patients counted multiply between the different analyses. |
| Treatment with PCSK9 monoclonal antibodies is associated with discontinuation of oral lipid lowering therapy | Engebretsen (2024)^51^ | 39562279 | Grace period | Both | 90 | No | Not calculable | Not calculable | 15%, 28%, 35% at 1/2/3 years' follow-up | Not calculable | Not calculable |  |
|  |  |  |  |  | 180 |  |  |  | 9%, 17%, 20% at 1/2/3 years' follow-up |  |  |  |
| Treatment adherence, persistence, and effectiveness of fixed dose combination versus free combination therapy of rosuvastatin–ezetimibe as a lipid-lowering therapy | Samnaliev (2025)^52^ | 40454236 | Maximum gap | Not specified | 45 | No | 7,642 | 2,866 | 37.50 | Not calculable | Not calculable | Computed mean/SD of age and numbers/proportions of males and discontinuers. |
| Effect of single-pill versus free equivalent combinations on persistence and major adverse cardiovascular events in hypertension: a real-world analysis | Simonyi (2025)^53^ | 39641301 | Grace period | Both | 90 | Yes | Not calculable | Not calculable | Not calculable | Not calculable | Not calculable | The authors describe trends in discontinuation over time, but no absolute numbers/proportions were reported. |

**Table S4 – Risk of bias of included studies.**

| **First Author (Year)** | **Associative measures?** | **Risk of bias due to confounding** | **Risk of bias arising from measurement of exposure** | **Risk of bias in selecting participants into the study** | **Risk of bias due to post-exposure interventions** | **Risk of bias due to missing data** | **Risk of bias arising from measurement of the outcome** | **Risk of bias in the selection of the reported result** | **Study-level RoB judgement** | **Notes** |
| --- | --- | --- | --- | --- | --- | --- | --- | --- | --- | --- |
| Ah (2015) | Age, sex | Some concerns | Low | Some concerns | Low | Low | Low | Low | Some concerns | Presents adjusted results without SEP/ethnicity adjustment. Participants were a randomly selected 20% of a fuller cohort of AHT initiators, but there is no comparison of the full and analysed cohorts in terms of age, sex, or comorbidity. |
| Mahmoudpour (2015) |  | Low | Low | Low | Low | Low | Low | Low | Low |  |
| Simonyi (2015) |  | Low | Low | Some concerns | Low | Low | Low | Low | Low | Unable to ascertain from text the specific inclusion/exclusion criteria, though a new user design is mentioned. |
| Ah (2016) | Age, sex | Some concerns | Low | Low | Low | Low | Low | Low | Some concerns | Presents adjusted results without SEP/ethnicity adjustment. |
| Burke (2016) |  | Low | Low | Low | Low | Low | Low | Low | Low |  |
| García-Gil (2016) |  | Low | Low | Low | Low | Some concerns | Low | Low | Some concerns | Replace baseline and 1-year missing values for certain covariates using multiple imputations by chain equations. Sensitivity analyses showed dissimilarity between complete and imputed cohorts for age/sex. |
| Halava (2016) | Age, sex | Some concerns | Low | Some concerns | Low | Low | Low | Low | Some concerns | No adjustment for ethnicity. Sample based on completion of survey in public sector workers, so may be unrepresentative. |
| Lin (2016) |  | Low | Low | Low | Low | Some concerns | Low | Low | Some concerns | Final sample excluded all patients with 'invalid days supply' (~15% of the sample). No discussion of whether missingness related to outcomes/exposures. |
| Quek (2016) | Age, sex | **Very high risk** |  |  |  |  |  |  | **Very high** | Estimates for age and sex are unadjusted risk ratios, so at very high risk of confounding . |
| Schaffer (2016) |  | Low | Low | Some concerns | Low | Low | Low | Low | Some concerns | Used a 10% random sample of existing data with no way to check if representative of full population. |
| Simonyi (2016) |  | Low | Low | Low | Low | Low | Low | Low | Low | The study had no associative measures of relevance to the review, even if its reported associations were highly likely to be confounded. |
| Vinogradova (2016) | Age, sex, SEP, ethnicity | Low | Low | Low | Low | Some concerns | Low | Low | Some concerns | Imputed missing data, and discuss a sensitivity analysis in which they compare imputed data against complete cases - but not present in supplementary files. |
| Karlsson (2017) | Age, sex, SEP | Low | Low | Low | Low | Low | Low | Low | Low | The ethnicity measures are based on country of birth instead of self-reported ethnicity, so may misclassify ethnicity. |
|  | Ethnicity | Low | Some concerns | Low | Low | Low | Low | Low | Some concerns |  |
| Tung (2017) | Age, sex | Some concerns | Low | Low | Low | Low | Low | Low | Some concerns | Presents adjusted results without SEP/ethnicity adjustment. |
| Wawruch (2017) | Age, sex | Some concerns | Low | Low | Low | Low | Low | Low | Some concerns | Presents adjusted results without SEP/ethnicity adjustment. |
| Chidwick (2018) | Age, sex | Some concerns | Low | Low | Low | Low | Low | Low | Some concerns | The age/sex estimates are not adjusted for SEP/ethnicity so may be at risk of bias due to confounding. The SEP estimates are unadjusted HRs and at high risk of bias from confounding. |
|  | SEP | **Very high risk** | Low | Low | Low | Low | Low | Low | **Very high** |  |
| Kim (2018) |  | Low | Low | Low | Low | Low | Low | Low | Low |  |
| Mahmoudpour (2018) |  | Low | Low | Low | Low | Low | Low | Low | Low |  |
| Malo (2018) | Age | Some concerns | Low | Low | Low | Low | Low | Low | Some concerns | Estimates not adjusted for SEP or ethnicity. Conducted in fully healthy, male population. |
| Nagar (2018) |  | Low | Low | Low | Low | Low | Low | Low | Low |  |
| Ofori-Asenso (2018a) | Age, sex | Some concerns | Low | Some concerns | Low | Low | Low | Some concerns | Some concerns | No adjustment for SEP or ethnicity. Concessional beneficiaries only, so representative of 65% of Australians. Associative measures reported in text with no reporting of full model. |
| Ofori-Asenso (2018b) | Sex | Some concerns | Low | Some concerns | Low | Low | Low | Some concerns | Some concerns | No adjustment for SEP or ethnicity. Alzheimer's identified by medication records only. Associative measures reported in text with no reporting of full model. |
| Ofori-Asenso (2018c) | Age, sex | Some concerns | Low | Low | Low | Low | Low | Some concerns | Some concerns | No adjustment for SEP or ethnicity. Associative measures reported in text with no reporting of full model. |
| Verma (2018) |  | Low | Low | Low | Low | Low | Low | Low | Low |  |
| Holvast (2019) |  | Low | Low | Low | Low | Low | Low | Low | Low |  |
| Morotti (2019) |  | Low | Low | Low | Low | Low | Low | Low | Low | Based in a veteran (so very male) population, but this isn’t a bias per se. |
| Ofori-Asenso (2019) | Age, sex | Some concerns | Low | Some concerns | Low | Low | Low | Low | Some concerns | No adjustment for ethnicity; analyses stratified on concessional status (i.e. an SEP proxy). All comorbidities derived using Rx-Risk-V tool which may misclassify an unknown proportion of cases. |
| Chen (2020) |  | Low | Low | Low | Low | Low | Low | Low | Low |  |
| Rezende Macedo do Nascimento (2020) |  | Low | Low | Low | Low | Low | Low | Low | Low |  |
| Sigglekow (2020) | Age, sex, SEP, ethnicity | Low | Low | Low | Low | Low | Low | Low | Low |  |
| Crockett (2021) |  | Low | Low | Some concerns | Low | Low | Low | Low | Some concerns | The study is 4:1 frequency matched non-HIV controls:patients with HIV. The prevalence of HIV in this cohort is therefore very high. As HIV is associated with non-persistence, discontinuation estimates in this paper will be higher than in other studies. |
| Gao (2021) |  | Low | Low | Some concerns | Low | Low | Low | Low | Some concerns | Propensity score matching means reported discontinuation proportions may be unrepresentative of the wider Japanese population. |
| Hsu (2021) |  | Low | Low | Low | Low | Low | Low | Low | Low |  |
| Wake (2021) |  | Low | Low | Low | Low | Low | Low | Low | Low |  |
| Beall (2022) | Age, sex, SEP, ethnicity | Low | Low | Low | Low | Low | Low | Low | Low | A small amount of missing data, but unlikely to bias results. The 7-day treatment gap may inflate the apparent rate of discontinuation but should not do this differentially across age/sex/SEP/ethnicity categories. |
| Engebretsen (2022) |  | Low | Low | Low | Low | Low | Low | Low | Low |  |
| Talic (2022) |  | Some concerns | Low | Low | Low | Low | Low | Low | Some concerns | No adjustment for ethnicity or SEP. Note that associative measures from this paper are not included in the review due to potential issues in the supplementary data which we have not been able to clarify with the study authors. |
| Eastwood (2023) | Age, sex, SEP, ethnicity | Low | Low | Low | Low | Low | Low | Low | Low |  |
| Kostev (2023) |  | Low | Low | Low | Low | Low | Low | Low | Low |  |
| Muntner (2023) | Age, sex | Some concerns | Low | Low | Low | Low | Low | Low | Some concerns | Presents adjusted results without SEP/ethnicity adjustment. |
| Engebretsen (2024) | Age, sex | Some concerns | Low | Low | Low | Low | Low | Low | Some concerns | Presents adjusted results without SEP/ethnicity adjustment. |
| Samnaliev (2025) |  | Low | Low | Low | Low | Low | Low | Low | Low |  |
| Simonyi (2025) |  | Low | Low | Low | Low | Low | Low | Low | Low |  |

**Table S5 – Discontinuation method descriptions and usage by study.**

| **Method** | **Description** |
| --- | --- |
| Grace period (or ‘refill-gap method’) | Patients discontinue if they have gaps in medication supply of more than some number of days.^10,11,14,15,17–22,25–28,30–33,36–39,41,44–47,49–51,53^  Some authors also account for medication stockpiling, by adding unused supply from a previous prescription on to the end of the next prescription. |
| Maximum gap | Patients discontinue if the gap between the start date of two subsequent prescription is more than some number of days, irrespective of the days’ supply of each prescription.^23,48,52^ |
| Relative window | Identical to the ‘grace period’ approach, except the grace period is either multiple of either the previous prescription’s duration or the median prescription length for that individual.^29,34,35^ |
| Anniversary | Patients discontinue if they do not have drug supply at specific dates after initiation (e.g. 1 year after index date).^40^ |
| Pre-anniversary window | Patients discontinue if they have no relevant prescriptions in a period immediately prior to 1 year after the index date.^42^ |
| Post-follow up window | Patients discontinue if they have no relevant prescription in a time window after the end of study follow-up.^16^ |
| Single prescription only | Patients considered to discontinue if they had only one prescription in the first year of follow-up.^17^ |
| Not specified | Methodology not specified.^24^ |

**Table S6 – Discontinuation and restarting prevalence by study population and use of medication ‘switching’ as discontinuation.**

| **Study population** | **Measure** | **Median (IQR)** | **References** |
| --- | --- | --- | --- |
| **Lipid-lowering therapies^a^ - Medication ‘switch’ as discontinuation^b^** | | | |
| Overall | Discontinuation | 62.9 (40.0 – 75.6) | ^16,19,30,39,40,42,44^ |
|  | Restarting | 29.4 (14.9 – 50.3) | ^19,30,39,40^ |
| Primary prevention | Discontinuation | 65.4 (39.7 – 75.9) | ^16,30,39,40,44^ |
|  | Restarting | 16.5 (14.3 – 44.2) | ^30,39,40^ |
| Secondary prevention | Discontinuation | 56.0 (48.0 – 71.7) | ^19,30,39,40,44^ |
|  | Restarting | 28.5 (13.5 – 51.1) | ^19,30,39,40^ |
| Mixed prevention | Discontinuation | 69.9 (51.5 – 79.6) | ^30,33,39,40,40,42,44^ |
|  | Restarting | 15.9 (13.9 – 44.1) | ^30,39,40^ |
| **Lipid-lowering therapies^a^ - Medication ‘switch’ not included as discontinuation** | | | |
| Overall | Discontinuation | 43.4 (33.4 – 58.8) | ^18,19,22,23,25–27,29–33,38,39,41,43,44,47,51^ |
|  | Restarting | 57.9 (39.5 – 60.7) | ^19,22,29,30,33,39,47^ |
| Primary prevention | Discontinuation | 38.8 (29.8 – 51.8) | ^22,23,26,29,30,39,41,43,44^ |
|  | Restarting | 41.3 (22.2 – 61.4) | ^19,22,30,39^ |
| Secondary prevention | Discontinuation | 39.3 (32.9 – 41.5) | ^19,22,23,25,30,39,41,43,44^ |
|  | Restarting | 41.0 (19.2 - 64.5) | ^19,22,30,39^ |
| Mixed prevention | Discontinuation | 45.3 (34.8 – 59.0) | ^18,22,23,27,30–33,38,39,41,43,44,47,51^ |
|  | Restarting | 55.2 (23.8 – 60.4) | ^22,30,33,39,47^ |
| **Antihypertensives - Medication ‘switch’ as discontinuation** | | | |
| Overall | Discontinuation | 49.4 (25.8 – 59.8) | ^10,11,14,21,28,48,50,53^ |
|  | Restarting | 17.6 | ^28^ |
| Primary prevention | Discontinuation | 64.0 (56.7 – 64.2) | ^10,14,21^ |
|  | Restarting | - | - |
| Secondary prevention | Discontinuation | - | - |
|  | Restarting | - | - |
| Mixed prevention | Discontinuation | 25.8 (11.3 – 43.7) | ^11,28,48,50,53^ |
|  | Restarting | 17.6 (17.6 – 17.6) | ^28^ |
| **Antihypertensives - Medication ‘switch’ not included as discontinuation** | | | |
| Overall | Discontinuation | 41.3 (30.4 – 49.4) | ^10,11,14,20,28,45^ |
|  | Restarting | 27.9 | ^28^ |
| Primary prevention | Discontinuation | 42.7 (41.3 – 47.2) | ^10,14,45^ |
|  | Restarting | - | - |
| Secondary prevention | Discontinuation | - | - |
|  | Restarting | - | - |
| Mixed prevention | Discontinuation | 27.2 (26.1 – 43.7) | ^11,20,28^ |
|  | Restarting | 27.9 | ^28^ |

^a^The Engebretsen *et al*. (2024) results used here are for the proportion discontinuing at 1 year with a 90-day grace period.^51^ ^b^Rezende Macedo do Nascimento *et al*. (2020) measured discontinuation using both the grace period and anniversary window methods.^40^ Both measures were included when computing medians and IQRs for discontinuation; only the grace period approach had an associated restarting measure.

**Table S7 – Measures of association between sociodemographic categories and lipid-lowering therapy discontinuation.**

| **Study or subgroup** | **Prevention** | **Switch as discontinuation?** | **Variable** | **Group** | **Total** | **Persistent** | **Discontinued** | **Proportion discontinued (3 d.p.)** | **Risk difference (2 d.p.)** | **Univariate^a^** | **Multivariate** | |
| --- | --- | --- | --- | --- | --- | --- | --- | --- | --- | --- | --- | --- |
|  |  |  |  |  |  |  |  |  |  | **RR/HR/OR [95% CI]** | **Model description** | **aHR/aOR**  **[95% CI]** |
| Quek (2015) [All patients] | Secondary | No | Age group | 18 to 64 | 175,046 | 115,383 | 59,663 | 0.341 | - | 1.00 (reference) | - | - |
|  |  |  |  | ≥65 | 37,232 | 23,441 | 13,791 | 0.370 | 0.03 | 1.09 [1.07 - 1.10] |  |  |
|  |  | Yes | Age group | 18 to 64 | 175,046 | 75,399 | 99,647 | 0.569 | - | 1.00 (reference) | - | - |
|  |  |  |  | ≥65 | 37,232 | 18,026 | 19,206 | 0.516 | -0.05 | 0.91 [0.90 - 0.92] |  |  |
| Quek (2015) [CVE subcohort] | Secondary | No | Age group | 18 to 64 | 34,674 | 23,405 | 11,269 | 0.325 | - | 1.00 (reference) | - | - |
|  |  |  |  | ≥65 | 7,260 | 4,828 | 2,432 | 0.335 | 0.01 | 1.03 [0.99 - 1.07] |  |  |
|  |  | Yes | Age group | 18 to 64 | 34,674 | 32,242 | 2,432 | 0.070 | - | 1.00 (reference) | - | - |
|  |  |  |  | ≥65 | 7,260 | 3,579 | 3,681 | 0.507 | 0.44 | 7.23 [6.91 - 7.56] |  |  |
| Quek (2015) [CHD-RE subcohort] | Secondary | No | Age group | 18 to 64 | 140,372 | 81,978 | 58,394 | 0.416 | - | 1.00 (reference) | - | - |
|  |  |  |  | ≥65 | 29,972 | 18,613 | 11,359 | 0.379 | -0.04 | 0.91 [0.90 - 0.93] |  |  |
|  |  | Yes | Age group | 18 to 64 | 140,372 | 59,518 | 80,854 | 0.576 | - | 1.00 (reference) | - | - |
|  |  |  |  | ≥65 | 29,972 | 14,447 | 15,525 | 0.518 | -0.06 | 0.90 [0.89 - 0.91] |  |  |
| Halava (2016) | Both | No | Age group | 24 to 50 | 1971 | 1,694 | 277 | 0.141 | - | 1.00 (reference) | Education, marital status, suboptimal self-rated health, antidepressant use, cancer, vascular comorbidity, co-payment of first package, body mass index, smoking status, risky alcohol use, physical activity. | 1.00 (reference) |
|  |  |  |  | 51 to 50 | 4809 | 4,243 | 566 | 0.118 | -0.02 | 0.84 [0.73 - 0.96] |  | 0.85 [0.72 - 1.01] |
|  |  |  |  | 61 to 75 | 2505 | 2206 | 299 | 0.119 | -0.02 | 0.85 [0.73 - 0.99] |  | 0.81 [0.68 - 0.98] |
|  |  |  | Sex | Male | 2208 | 1,930 | 278 | 0.126 | - | 1.00 (reference) |  | 1.00 (reference) |
|  |  |  |  | Female | 7077 | 6,213 | 864 | 0.122 | 0.00 | 0.97 [0.86 - 1.10] |  | 1.01 [0.86 - 1.17] |
| Vinogradova (2016) | Primary | Not specified | Age group | 25 to 45 | 31,290 | 12,303 | 18,987 | 0.607 | - | 1.00 (reference) | Model includes smoking status, chronic conditions, use of other drugs, practice-based Townsend score groups, year of entering the study, body mass index as covariates. Age was included as a fractional polynomial so is not available on a category-by-category basis. More details are available in the Table 3 legend in the paper. | - |
|  |  |  |  | 45 to 54 | 77,770 | 37,191 | 40,579 | 0.522 | -0.09 | 0.86 [0.85 - 0.87] |  |  |
|  |  |  |  | 55 to 64 | 140,042 | 76,395 | 63,647 | 0.454 | -0.15 | 0.75 [0.74 - 0.76] |  |  |
|  |  |  |  | 65 to 74 | 129,429 | 72,574 | 56,855 | 0.439 | -0.17 | 0.72 [0.72 - 0.73] |  |  |
|  |  |  |  | 75 to 84 | 52,492 | 27,938 | 24,554 | 0.468 | -0.14 | 0.77 [0.76 - 0.78] |  |  |
|  |  |  | Sex | Male | 225,732 | 120,423 | 105,309 | 0.467 | - | 1.00 (reference) |  | 1.00 (reference) |
|  |  |  |  | Female | 205,291 | 105,978 | 99,313 | 0.484 | 0.02 | 1.04 [1.03 - 1.04] |  | 1.06 [1.05 - 1.07] |
|  |  |  | SEP (Patient-based Townsend score quintile) | 5^th^ quintile (low) | 32,902 | 16,567 | 16,335 | 0.496 | - | 1.00 (reference) |  | 0.98 [0.95 - 1.02] |
|  |  |  |  | 4^th^ quintile | 50,631 | 26,496 | 24,135 | 0.477 | -0.02 | 0.96 [0.95 - 0.97] |  | 0.99 [0.96 - 1.02] |
|  |  |  |  | 3^rd^ quintile | 54,715 | 28,522 | 26,193 | 0.479 | -0.02 | 0.96 [0.95 - 0.98] |  | 1.02 [1.00 - 1.05] |
|  |  |  |  | 2^nd^ quintile | 62,581 | 33,141 | 29,440 | 0.470 | -0.03 | 0.95 [0.93 - 0.96] |  | 1.02 [1.00 - 1.04] |
|  |  |  |  | 1^st^ quintile (high) | 65,226 | 34,761 | 30,465 | 0.467 | -0.03 | 0.94 [0.93 - 0.95] |  | 1.00 (reference) |
|  |  |  | SEP (Practice-based Townsend score quintile) | 5^th^ quintile (low) | 89,653 | 47,474 | 42,179 | 0.470 | - | 1.00 (reference) |  | - |
|  |  |  |  | 4^th^ quintile | 94,933 | 49,729 | 45,204 | 0.476 | 0.01 | 1.01 [1.00 - 1.02] |  |  |
|  |  |  |  | 3^rd^ quintile | 87,529 | 46,375 | 41,154 | 0.470 | 0.00 | 1.00 [0.99 - 1.01] |  |  |
|  |  |  |  | 2^nd^ quintile | 81,744 | 42,737 | 39,007 | 0.477 | 0.01 | 1.01 [1.00 - 1.02] |  |  |
|  |  |  |  | 1^st^ quintile (high) | 77,164 | 40,086 | 37,078 | 0.481 | 0.01 | 1.02 [1.01 - 1.03] |  |  |
|  |  |  | Ethnicity | White/Not recorded | 415,392 | 220,234 | 195,158 | 0.470 | - | 1.00 (reference) |  | 1.00 (reference) |
|  |  |  |  | Indian | 4,306 | 1,730 | 2,576 | 0.598 | 0.13 | 1.27 [1.24 - 1.31] |  | 1.45 [1.39 - 1.52] |
|  |  |  |  | Pakistani | 1,786 | 672 | 1,114 | 0.624 | 0.15 | 1.33 [1.28 - 1.38] |  | 1.48 [1.39 - 1.58] |
|  |  |  |  | Bangladeshi | 642 | 265 | 377 | 0.587 | 0.12 | 1.25 [1.17 - 1.33] |  | 1.18 [0.99 - 1.41] |
|  |  |  |  | Other Asian | 1,881 | 783 | 1,098 | 0.584 | 0.11 | 1.24 [1.20 - 1.29] |  | 1.37 [1.26 - 1.49] |
|  |  |  |  | Black African | 1,358 | 457 | 901 | 0.663 | 0.19 | 1.41 [1.36 - 1.47] |  | 1.89 [1.72 - 2.08] |
|  |  |  |  | Black Carribbean | 1,882 | 633 | 1,249 | 0.664 | 0.19 | 1.41 [1.37 - 1.46] |  | 1.89 [1.74 - 2.04] |
|  |  |  |  | Chinese | 619 | 283 | 336 | 0.543 | 0.07 | 1.16 [1.07 - 1.24] |  | 1.25 [1.11 - 1.41] |
|  | Secondary | Not specified | Age group | 25 to 45 | 4,158 | 2,167 | 1,991 | 0.479 | - | 1.00 (reference) | Model includes smoking status, chronic conditions, use of other drugs, practice-based Townsend score groups, year of entering the study, body mass index as covariates. Age was included as a fractional polynomial so is not available on a category-by-category basis. More details are available in the Table 4 legend in the paper. | - |
|  |  |  |  | 45 to 54 | 15,051 | 8,923 | 6,128 | 0.407 | -0.07 | 0.85 [0.82 - 0.88] |  |  |
|  |  |  |  | 55 to 64 | 32,620 | 19,995 | 12,625 | 0.387 | -0.09 | 0.81 [0.78 - 0.84] |  |  |
|  |  |  |  | 65 to 74 | 44,055 | 26,113 | 17,942 | 0.407 | -0.07 | 0.85 [0.82 - 0.88] |  |  |
|  |  |  |  | 75 to 84 | 43,430 | 24,325 | 19,105 | 0.440 | -0.04 | 0.92 [0.89 - 0.95] |  |  |
|  |  |  | Sex | Male | 82,485 | 50,353 | 32,132 | 0.390 | - | 1.00 (reference) |  | 1.00 (reference) |
|  |  |  |  | Female | 56,829 | 31,170 | 25,659 | 0.452 | 0.06 | 1.16 [1.14 - 1.17] |  | 1.14 [1.12 - 1.17] |
|  |  |  | SEP (Patient-based Townsend score) | 5^th^ quintile (low) | 10,988 | 6,333 | 4,655 | 0.424 | - | 1.00 (reference) |  | 0.97 [0.92 - 1.01] |
|  |  |  |  | 4^th^ quintile | 16,898 | 9,639 | 7,259 | 0.430 | 0.01 | 1.01 [0.99 - 1.04] |  | 1.02 [0.97 - 1.06] |
|  |  |  |  | 3^rd^ quintile | 17,905 | 10,544 | 7,361 | 0.411 | -0.01 | 0.97 [0.94 - 1.00] |  | 0.99 [0.95 - 1.03] |
|  |  |  |  | 2^nd^ quintile | 19,647 | 11,491 | 8,156 | 0.415 | -0.01 | 0.98 [0.95 - 1.01] |  | 1.02 [0.98 - 1.06] |
|  |  |  |  | 1^st^ quintile (high) | 18,966 | 11,286 | 7,680 | 0.405 | -0.02 | 0.96 [0.93 - 0.98] |  | 1.00 (reference) |
|  |  |  | SEP (Practice-based Townsend score) | 5^th^ quintile (low) | 22,887 | 13,399 | 9,488 | 0.415 | - | 1.00 (reference) |  | - |
|  |  |  |  | 4^th^ quintile | 25,379 | 14,988 | 10,391 | 0.409 | -0.01 | 0.99 [0.97 - 1.01] |  |  |
|  |  |  |  | 3^rd^ quintile | 28,498 | 16,669 | 11,829 | 0.415 | 0.00 | 1.00 [0.98 - 1.02] |  |  |
|  |  |  |  | 2^nd^ quintile | 32,131 | 18,752 | 13,379 | 0.416 | 0.00 | 1.00 [0.98 - 1.02] |  |  |
|  |  |  |  | 1^st^ quintile (high) | 30,419 | 17,715 | 12,704 | 0.418 | 0.00 | 1.01 [0.99 - 1.03] |  |  |
|  |  |  | Ethnicity | White/Not recorded | 136,259 | 79,986 | 56,273 | 0.413 | - | 1.00 (reference) |  | 1.00 (reference) |
|  |  |  |  | Indian | 953 | 479 | 474 | 0.497 | 0.08 | 1.20 [1.13 - 1.28] |  | 1.35 [1.23 - 1.49] |
|  |  |  |  | Pakistani | 422 | 218 | 204 | 0.483 | 0.07 | 1.17 [1.06 - 1.29] |  | 1.41 [1.19 - 1.66] |
|  |  |  |  | Bangladeshi | 121 | 67 | 54 | 0.446 | 0.03 | 1.08 [0.89 - 1.32] |  | 1.17 [0.84 - 1.61] |
|  |  |  |  | Other Asian | 229 | 106 | 123 | 0.537 | 0.12 | 1.30 [1.15 - 1.47] |  | 1.52 [1.30 - 1.79] |
|  |  |  |  | Black African | 165 | 79 | 86 | 0.521 | 0.11 | 1.26 [1.09 - 1.46] |  | 1.69 [1.33 - 2.16] |
|  |  |  |  | Black Carribbean | 368 | 146 | 222 | 0.603 | 0.19 | 1.46 [1.34 - 1.59] |  | 1.80 [1.54 - 2.10] |
|  |  |  |  | Chinese | 123 | 71 | 52 | 0.423 | 0.01 | 1.02 [0.83 - 1.26] |  | 1.17 [0.87 - 1.57] |
|  | Both | Not specified | Age group | 25 to 45 | 35,448 | 14,470 | 20,978 | 0.592 | - | 1.00 (reference) | - | |
|  |  |  |  | 45 to 54 | 92,821 | 46,114 | 46,707 | 0.503 | -0.09 | 0.85 [0.84 - 0.86] |  |  |
|  |  |  |  | 55 to 64 | 172,662 | 96,390 | 76,272 | 0.442 | -0.15 | 0.75 [0.74 - 0.75] |  |  |
|  |  |  |  | 65 to 74 | 173,484 | 98,687 | 74,797 | 0.431 | -0.16 | 0.73 [0.72 - 0.74] |  |  |
|  |  |  |  | 75 to 84 | 95,922 | 52,263 | 43,659 | 0.455 | -0.14 | 0.77 [0.76 - 0.78] |  |  |
|  |  |  | Sex | Male | 308,217 | 170,776 | 137,441 | 0.446 | - | 1.00 (reference) |  |  |
|  |  |  |  | Female | 262,120 | 137,148 | 124,972 | 0.477 | 0.03 | 1.07 [1.06 - 1.08] |  |  |
|  |  |  | SEP (Patient-based Townsend score) | 5^th^ quintile (low) | 43,890 | 22,900 | 20,990 | 0.478 | - | 1.00 (reference) |  |  |
|  |  |  |  | 4^th^ quintile | 67,529 | 36,135 | 31,394 | 0.465 | -0.01 | 0.97 [0.96 - 0.98] |  |  |
|  |  |  |  | 3^rd^ quintile | 72,620 | 39,066 | 33,554 | 0.462 | -0.02 | 0.97 [0.95 - 0.98] |  |  |
|  |  |  |  | 2^nd^ quintile | 82,228 | 44,632 | 37,596 | 0.457 | -0.02 | 0.96 [0.94 - 0.97] |  |  |
|  |  |  |  | 1^st^ quintile (high) | 84,192 | 46,047 | 38,145 | 0.453 | -0.03 | 0.95 [0.94 - 0.96] |  |  |
|  |  |  | SEP (Practice-based Townsend score) | 5^th^ quintile (low) | 120,072 | 65,189 | 54,883 | 0.457 | - | 1.00 (reference) |  |  |
|  |  |  |  | 4^th^ quintile | 127,064 | 68,481 | 58,583 | 0.461 | 0.00 | 1.01 [1.00 - 1.02] |  |  |
|  |  |  |  | 3^rd^ quintile | 116,027 | 63,044 | 52,983 | 0.457 | 0.00 | 1.00 [0.99 - 1.01] |  |  |
|  |  |  |  | 2^nd^ quintile | 107,123 | 57,725 | 49,398 | 0.461 | 0.00 | 1.01 [1.00 - 1.02] |  |  |
|  |  |  |  | 1^st^ quintile (high) | 100,051 | 53,485 | 46,566 | 0.465 | 0.01 | 1.02 [1.01 - 1.03] |  |  |
|  |  |  | Ethnicity | White/Not recorded | 551,651 | 300,220 | 251,431 | 0.456 | - | 1.00 (reference) |  |  |
|  |  |  |  | Indian | 5,259 | 2,209 | 3,050 | 0.580 | 0.12 | 1.27 [1.24 - 1.30] |  |  |
|  |  |  |  | Pakistani | 2,208 | 890 | 1,318 | 0.597 | 0.14 | 1.31 [1.27 - 1.36] |  |  |
|  |  |  |  | Bangladeshi | 763 | 332 | 431 | 0.565 | 0.11 | 1.24 [1.16 - 1.32] |  |  |
|  |  |  |  | Other Asian | 2,110 | 889 | 1,221 | 0.579 | 0.12 | 1.27 [1.22 - 1.32] |  |  |
|  |  |  |  | Black African | 1,523 | 536 | 987 | 0.648 | 0.19 | 1.42 [1.37 - 1.48] |  |  |
|  |  |  |  | Black Carribbean | 2,250 | 779 | 1,471 | 0.654 | 0.20 | 1.43 [1.39 - 1.48] |  |  |
|  |  |  |  | Chinese | 742 | 354 | 388 | 0.523 | 0.07 | 1.15 [1.07 - 1.23] |  |  |
| Wawruch (2016) [Excluding patients with stroke/MI in follow-up] | Secondary | No | Age group | 65 to 74 | 1,312 | 695 | 617 | 0.470 | - | 1.00 (reference) | Both models also include arterial hypertension, diabetes mellitus, hypercholesterolemia, dementia, depression, anxiety disorders, Parkinson’s disease, epilepsy, and polypharmacy (receiving ≥6 drugs). | 1.00 (reference) |
|  |  |  |  | ≥75 | 1,436 | 961 | 475 | 0.331 | -0.14 | 0.70 [0.64 - 0.77] |  | 0.75 [0.67 - 0.85] |
|  |  |  | Sex | Male | 1,150 | 699 | 451 | 0.392 | - | 1.00 (reference) |  | 1.00 (reference) |
|  |  |  |  | Female | 1,598 | 957 | 641 | 0.401 | 0.01 | 1.02 [0.93 - 1.12] |  | 1.04 [0.92 - 1.18] |
| Wawruch (2016) [Including patients with stroke/MI in follow-up] | Secondary | No | Age group | 65 to 74 | 1,635 | 1,018 | 617 | 0.377 | - | 1.00 (reference) |  | 1.00 (reference) |
|  |  |  |  | ≥75 | 1,809 | 1,334 | 475 | 0.263 | -0.11 | 0.70 [0.63 - 0.77] |  | 0.74 [0.65 - 0.83] |
|  |  |  | Sex | Male | 1,421 | 970 | 451 | 0.317 | - | 1.00 (reference) |  | 1.00 (reference) |
|  |  |  |  | Female | 2,023 | 1,382 | 641 | 0.317 | 0.00 | 1.00 [0.90 - 1.10] |  | 1.08 [0.95 - 1.22] |
| Karlsson (2017) | Both | No | Age | - | - | | | | | | Model includes marital status, education level, employment status, anticoagulant use, diabetes medication use, antihypertensive use, low- and high-density lipoprotein cholesterol, total cholesterol, blood triglycerides, HbA1c, estimated glomerular filtration rate, body mass index, systolic/diastolic blood pressure, micro- or macroalbuminuria, cancer diagnosis, physical activity, and smoking status. Age was included as a continuous covariate so is not available on a category-by-category basis. | 1.00 [0.99 - 1.00] |
|  |  |  | Sex | Male |  |  |  |  |  |  |  | 1.00 (reference) |
|  |  |  |  | Female |  |  |  |  |  |  |  | 1.11 [1.05 - 1.16] |
|  |  |  | SEP (individual disposable income quartile) | 1^st^ quartile (low) |  |  |  |  |  |  |  | 1.00 (reference) |
|  |  |  |  | 2^nd^ quartile |  |  |  |  |  |  |  | 0.86 [0.80 - 0.93] |
|  |  |  |  | 3^rd^ quartile |  |  |  |  |  |  |  | 0.87 [0.81 - 0.93] |
|  |  |  |  | 4^th^ quartile (high) |  |  |  |  |  |  |  | 0.84 [0.77 - 0.91] |
|  |  |  | SEP (occupation) | Blue collar |  |  |  |  |  |  |  | 1.00 (reference) |
|  |  |  |  | Lower white collar |  |  |  |  |  |  |  | 1.03 [0.97 - 1.10] |
|  |  |  |  | Upper white collar |  |  |  |  |  |  |  | 1.01 [0.94 - 1.10] |
|  |  |  |  | Other |  |  |  |  |  |  |  | 0.99 [0.87 - 1.12] |
|  |  |  | Ethnicity (proxied by country of birth) | Sweden |  |  |  |  |  |  |  | 1.00 (reference) |
|  |  |  |  | Other Nordic country |  |  |  |  |  |  |  | 1.16 [1.05 - 1.28] |
|  |  |  |  | Other EU27 country |  |  |  |  |  |  |  | 1.19 [1.03 - 1.38] |
|  |  |  |  | Rest of Europe/the Soviet Union |  |  |  |  |  |  |  | 1.25 [1.07 - 1.47] |
|  |  |  |  | The Americas |  |  |  |  |  |  |  | 1.64 [1.33 - 2.02] |
|  |  |  |  | Asia/Oceania |  |  |  |  |  |  |  | 1.81 [1.44 - 2.27] |
| Malo (2017) | Primary | No | Age group | <50 | 95 | - | | | | 1.00 (reference) | Antidiabetic use, antihypertensive use, antithrombotic use, baseline LDL-C, baseline total cholesterol. | 1.00 (reference) |
|  |  |  |  | 50 to 54 | 231 |  |  |  |  | 0.95 [0.72 - 1.24] |  | 0.98 [0.74 - 1.31] |
|  |  |  |  | 55 to 59 | 267 |  |  |  |  | 0.73 [0.56 - 0.96] |  | 0.79 [0.59 - 1.05] |
|  |  |  |  | ≥60 | 132 |  |  |  |  | 0.49 [0.35 - 0.68] |  | 0.55 [0.39 - 0.77] |
| Chidwick (2018)^c^ | Primary | No | Age group | 40 to 49 | 44,088 | 17,635 | 26,453 | 0.600 | - | 1.00 (reference) | Main exposure was cancer survivor status. Model also includes body mass index, smoking status and diabetes status (time-updated). | 1.00 (reference) |
|  |  |  |  | 50 to 59 | 95,539 | 44,440 | 51,099 | 0.535 | -0.07 | 0.81 [0.80 - 0.82] |  | 0.80 [0.79 - 0.82] |
|  |  |  |  | 60 to 69 | 126,570 | 65,807 | 60,763 | 0.480 | -0.12 | 0.69 [0.68 - 0.70] |  | 0.66 [0.65 - 0.67] |
|  |  |  |  | 70 to 79 | 75,425 | 37,778 | 37,647 | 0.499 | -0.10 | 0.73 [0.71 - 0.75] |  | 0.68 [0.67 - 0.70] |
|  |  |  |  | ≥80 | 25,149 | 11,305 | 13,844 | 0.550 | -0.05 | 0.89 [0.85 - 0.93] |  | 1.14 [1.05 - 1.25] |
|  |  |  | Sex | Male | 193,646 | 95,400 | 98,246 | 0.507 | - | 1.00 (reference) |  | 1.00 (reference) |
|  |  |  |  | Female | 173,125 | 81,565 | 91,560 | 0.529 | 0.02 | 1.05 [1.04 - 1.06] |  | 1.05 [1.04 - 1.06] |
|  |  |  | SEP (IMD quintiles) | Least deprived quintile | 40,769 | 19,717 | 21,052 | 0.516 | - | 1.00 (reference) |  | - |
|  |  |  |  | Second-least deprived quintile | 43,256 | 21,159 | 22,097 | 0.511 | 0.00 | 1.01 [0.97 - 1.06] |  |  |
|  |  |  |  | Middle quintile | 48,504 | 23,596 | 24,908 | 0.514 | 0.00 | 1.03 [0.98 - 1.09] |  |  |
|  |  |  |  | Second-most deprived quintile | 187,806 | 89,776 | 98,030 | 0.522 | 0.01 | 1.11 [1.06 - 1.17] |  |  |
|  |  |  |  | Most deprived quintile | 46,436 | 22,717 | 23,719 | 0.511 | 0.00 | 1.03 [0.97 - 1.09] |  |  |
| Ofori-Asenso (2018a) ["Patterns of statin use..."] | Both | No | Age group | 65 to 74 | - | | | | | | Statin type, statin intensity, prescriber type, and initiation year. | 1.00 (reference) |
|  |  |  |  | 75 to 84 |  |  |  |  |  |  |  | 1.03 [0.97 - 1.10] |
|  |  |  |  | ≥85 |  |  |  |  |  |  |  | 0.91 [0.81 - 1.02] |
|  |  |  | Sex | Male | 3,981 | - | | | | |  | 1.00 (reference) |
|  |  |  |  | Female | 3,419 |  |  |  |  |  |  | 1.06 [0.99 – 1.12] |
| Ofori-Asenso (2018b) ["Prevalence and incidence..."] | Both | No | Sex | Male | 212 | - | | | | | Age, statin type, intensity, and status of dementia medication use. | 1.00 (reference) |
|  |  |  |  | Female | 377 |  |  |  |  |  |  | 0.80 [0.64 - 1.01] |
| Ofori-Asenso (2018c) ["Switching, Discontinuation..."] | Both | No | Age group | 65 to 74 | 31,105 | - | | | | |  | 1.00 (reference) |
|  |  |  |  | 75 to 84 | 14,179 |  |  |  |  |  |  | 1.06 [1.03 - 1.09] |
|  |  |  |  | ≥85 | 4,096 |  |  |  |  |  |  | 1.14 [1.09 - 1.19] |
|  |  |  | Sex | Male | 22,585 | - | | | | |  | 1.00 (reference) |
|  |  |  |  | Female | 26,795 |  |  |  |  |  |  | 1.05 [1.03 – 1.08] |
| Ofori-Asenso (2019) [All patients] | Both | No | Age group | 65 to 74 | 14,332 | 7,910 | 6,422 | 0.448 | - | 1.00 (reference) | - | |
|  |  |  |  | 75 to 84 | 6,183 | 3,501 | 2,682 | 0.434 | -0.01 | 0.97 [0.94 - 1.00] |  |  |
|  |  |  |  | ≥85 | 1,825 | 953 | 872 | 0.478 | 0.03 | 1.07 [1.01 - 1.12] |  |  |
|  |  |  | Sex | Male | 10,828 | 5,922 | 4,906 | 0.453 | - | 1.00 (reference) |  |  |
|  |  |  |  | Female | 11,512 | 6,440 | 5,072 | 0.441 | -0.01 | 0.97 [0.94 - 1.00] |  |  |
| Ofori-Asenso (2019)^d^ [Concessional beneficiaries] | Both | No | Age group | 65 to 74 | 10,345 | 5,897 | 4,448 | 0.430 | - | 1.00 (reference) | Statin type, index prescriber, comorbidities (diabetes mellitus, angina, anxiety, steroid-responsive condition, hypertension, congestive heart failure, pain), use of platelet inhibitors, use of anticoagulant therapy, polypharmacy (≥ 5 medications). | 1.00 (reference) |
|  |  |  |  | 75 to 84 | 5,510 | 3,174 | 2,336 | 0.424 | -0.01 | 0.99 [0.95 - 1.02] |  | 1.03 [0.96 - 1.10] |
|  |  |  |  | ≥85 | 1,644 | 891 | 753 | 0.458 | 0.03 | 1.07 [1.01 - 1.13] |  | 1.19 [1.07 - 1.33] |
|  |  |  | Sex | Male | 9,053 | 5,052 | 4,001 | 0.442 | - | 1.00 (reference) |  | - |
|  |  |  |  | Female | 8,446 | 4,907 | 3,539 | 0.419 | -0.02 | 0.95 [0.91 - 0.98] |  |  |
| Ofori-Asenso (2019)^d^ [General beneficiaries] | Both | No | Age group | 65 to 74 | 3,987 | 2,013 | 1,974 | 0.495 | - | 1.00 (reference) | Statin type, index prescriber, comorbidities (diabetes mellitus, angina, anxiety, steroid-responsive condition, hypertension, congestive heart failure, pain), use of platelet inhibitors, use of anticoagulant therapy, polypharmacy (≥ 5 medications). | 1.00 (reference) |
|  |  |  |  | 75 to 84 | 673 | 327 | 346 | 0.514 | 0.02 | 1.04 [0.96 - 1.12] |  | 1.08 [0.92 - 1.28] |
|  |  |  |  | ≥85 | 181 | 62 | 119 | 0.657 | 0.16 | 1.33 [1.19 - 1.48] |  | 1.99 [1.45 - 2.75] |
|  |  |  | Sex | Male | 1,775 | 870 | 905 | 0.510 | - | 1.00 (reference) |  | - |
|  |  |  |  | Female | 3,066 | 1,533 | 1,533 | 0.500 | -0.01 | 0.98 [0.93 - 1.04] |  |  |
| Sigglekow (2020)^e^ | Primary | No | Age at first dispensing | <35 | 6,165 | 2,978 | 3,187 | 0.517 | - | 2.86 [2.72 - 3.02] | Modified Charlson comorbidity index score, year of first statin dispensing, scope of practice of first statin prescriber, first statin dispensed, defined daily dose of statin, and days’ supply of first statin dispensation. | 2.48 [2.36 - 2.62] |
|  |  |  |  | 35 to 44 | 24,009 | 14,045 | 9,964 | 0.415 | -0.10 | 1.90 [1.84 - 1.96] |  | 1.71 [1.66 - 1.76] |
|  |  |  |  | 45 to 54 | 58,069 | 37,919 | 20,150 | 0.347 | -0.17 | 1.42 [1.38 - 1.45] |  | 1.35 [1.31 - 1.38] |
|  |  |  |  | 55 to 64 | 75,062 | 54,645 | 20,417 | 0.272 | -0.24 | 1.00 (reference) |  | 1.00 (reference) |
|  |  |  |  | 65 to 74 | 51,705 | 39,606 | 12,099 | 0.234 | -0.28 | 0.81 [0.79 - 0.84] |  | 0.83 [0.81 - 0.85] |
|  |  |  |  | ≥75 | 23,845 | 18,551 | 5,294 | 0.222 | -0.29 | 0.76 [0.74 - 0.79] |  | 0.80 [0.77 - 0.83] |
|  |  |  | Sex | Male | 128,088 | 88,893 | 39,195 | 0.306 | - | 1.00 (reference) |  | 1.00 (reference) |
|  |  |  |  | Female | 110,740 | 78,847 | 31,893 | 0.288 | -0.02 | 0.91 [0.90 - 0.93] |  | 0.97 [0.95 - 0.99] |
|  |  |  |  | Unspecified | 27 | 22 | 5 | 0.185 | -0.12 | 0.51 [0.17 - 1.25] |  | 0.47 [0.16 - 1.17] |
|  |  |  | SEP (NZdep06 quintile) | 5^th^ quintile (low) | 59,215 | 44,530 | 14,685 | 0.248 | - | 1.00 (reference) |  | 1.11 [1.08 - 1.15] |
|  |  |  |  | 4^th^ quintile | 50,701 | 39,952 | 10,749 | 0.212 | -0.04 | 0.86 [0.83 - 0.87] |  | 1.03 [0.99 - 1.06] |
|  |  |  |  | 3^rd^ quintile | 44,643 | 36,295 | 8,348 | 0.187 | -0.06 | 0.75 [0.74 - 0.77] |  | 1.03 [1.00 - 1.07] |
|  |  |  |  | 2^nd^ quintile | 35,360 | 30,127 | 5,233 | 0.148 | -0.10 | 0.60 [0.58 - 0.61] |  | 1.02 [0.99 - 1.06] |
|  |  |  |  | 1^st^ quintile (high) | 33,359 | 28,689 | 4,670 | 0.140 | -0.11 | 0.56 [0.55 - 0.58] |  | 1.00 (reference) |
|  |  |  |  | Unspecified | 15,577 | 14,564 | 1,013 | 0.065 | -0.18 | 0.26 [0.25 - 0.28] |  | 1.06 [1.01 - 1.10] |
|  |  |  | Ethnicity (prioritised) | European | 143,107 | 105,470 | 37,637 | 0.263 | - | 1.00 (reference) |  | 1.00 (reference) |
|  |  |  |  | Maori | 20,367 | 12,505 | 7,862 | 0.386 | 0.12 | 1.76 [1.71 - 1.81] |  | 1.48 [1.43 - 1.53] |
|  |  |  |  | Pacific Peoples | 14,461 | 7,968 | 6,493 | 0.449 | 0.19 | 2.28 [2.20 - 2.36] |  | 1.84 [1.78 - 1.91] |
|  |  |  |  | Asian | 19,840 | 12,519 | 7,321 | 0.369 | 0.11 | 1.63 [1.58 - 1.69] |  | 1.42 [1.37 - 1.46] |
|  |  |  |  | MELAA | 16,291 | 11,567 | 4,724 | 0.290 | 0.03 | 1.14 [1.10 - 1.18] |  | 1.08 [1.05 - 1.13] |
|  |  |  |  | Other | 99 | 76 | 23 | 0.232 | -0.03 | 0.85 [0.52 - 1.33] |  | 0.80 [0.49 - 1.25] |
|  |  |  |  | Unknown | 24,690 | 17,678 | 7,012 | 0.284 | 0.02 | 1.11 [1.08 - 1.15] |  | 1.08 [1.05 - 1.11] |
|  | Secondary | No | Age group | <35 | 251 | 156 | 95 | 0.378 | - | 2.46 [1.90 - 3.18] | Modified Charlson comorbidity index score, year of first statin dispensing, scope of practice of first statin prescriber, first statin dispensed, defined daily dose of statin, and days’ supply of first statin dispensation. | 2.32 [1.83 - 3.00] |
|  |  |  |  | 35 to 44 | 2,275 | 1,661 | 614 | 0.270 | -0.11 | 1.49 [1.35 - 1.66] |  | 1.47 [1.32 - 1.63] |
|  |  |  |  | 45 to 54 | 7,470 | 5,782 | 1,688 | 0.226 | -0.15 | 1.18 [1.10 - 1.27] |  | 1.17 [1.09 - 1.25] |
|  |  |  |  | 55 to 64 | 12,228 | 9,807 | 2,421 | 0.198 | -0.18 | 1.00 (reference) |  | 1.00 (reference) |
|  |  |  |  | 65 to 74 | 12,810 | 10,504 | 2,306 | 0.180 | -0.20 | 0.89 [0.83 - 0.95] |  | 0.88 [0.82 - 0.94] |
|  |  |  |  | ≥75 | 15,777 | 12,874 | 2,903 | 0.184 | -0.19 | 0.91 [0.86 - 0.97] |  | 0.90 [0.84 - 0.96] |
|  |  |  | Sex | Male | 30,705 | 25,055 | 5,650 | 0.184 | - | 1.00 (reference) |  | 1.00 (reference) |
|  |  |  |  | Female | 20,105 | 15,722 | 4,383 | 0.218 | 0.03 | 1.24 [1.19 - 1.30] |  | 1.23 [1.17 - 1.28] |
|  |  |  | SEP (NZdep06 quintile) | 5^th^ quintile (low) | 12,719 | 9,539 | 3,180 | 0.250 | - | 1.00 (reference) |  | 1.11 [1.02 - 1.20] |
|  |  |  |  | 4^th^ quintile | 11,680 | 8,994 | 2,686 | 0.230 | -0.02 | 0.92 [0.88 - 0.96] |  | 0.99 [0.92 - 1.08] |
|  |  |  |  | 3^rd^ quintile | 9,624 | 7,805 | 1,819 | 0.189 | -0.06 | 0.76 [0.72 - 0.80] |  | 1.02 [0.94 - 1.09] |
|  |  |  |  | 2^nd^ quintile | 6,931 | 5,988 | 943 | 0.136 | -0.11 | 0.54 [0.51 - 0.58] |  | 1.00 [0.92 - 1.09] |
|  |  |  |  | 1^st^ quintile (high) | 6,407 | 5,600 | 807 | 0.126 | -0.12 | 0.50 [0.47 - 0.54] |  | 1.00 (reference) |
|  |  |  |  | Unspecified | 3,450 | 3,215 | 235 | 0.068 | -0.18 | 0.27 [0.24 - 0.31] |  | 1.01 [0.91 - 1.13] |
|  |  |  | Ethnicity (prioritised) | European | 38,646 | 31,535 | 7,111 | 0.184 | - | 1.00 (reference) |  | 1.00 (reference) |
|  |  |  |  | Maori | 4,762 | 3,476 | 1,286 | 0.270 | 0.09 | 1.64 [1.53 - 1.76] |  | 1.45 [1.33 - 1.56] |
|  |  |  |  | Pacific Peoples | 1,968 | 1,380 | 588 | 0.299 | 0.11 | 1.89 [1.71 - 2.09] |  | 1.71 [1.54 - 1.90] |
|  |  |  |  | Asian | 1,823 | 1,389 | 434 | 0.238 | 0.05 | 1.38 [1.24 - 1.52] |  | 1.30 [1.19 - 1.49] |
|  |  |  |  | MELAA | 1,611 | 1,339 | 272 | 0.169 | -0.02 | 0.90 [0.79 - 1.03] |  | 0.90 [0.78 - 1.02] |
|  |  |  |  | Other | 14 | 13 | 1 | 0.071 | -0.11 | 0.34 [0.02 - 1.71] |  | 0.29 [0.02 -1.47] |
|  |  |  |  | Unknown | 1,987 | 1,641 | 346 | 0.174 | -0.01 | 0.93 [0.83 - 1.05] |  | 0.96 [0.85 - 1.08] |
|  | Both | No | Age group | <35 | 6,416 | 3,134 | 3,282 | 0.512 | - | 1.00 (reference) | - | |
|  |  |  |  | 35 to 44 | 26,284 | 15,706 | 10,578 | 0.402 | -0.11 | 0.78 [0.76 - 0.81] |  |  |
|  |  |  |  | 45 to 54 | 65,539 | 43,701 | 21,838 | 0.333 | -0.18 | 0.65 [0.64 - 0.67] |  |  |
|  |  |  |  | 55 to 64 | 87,290 | 64,452 | 22,838 | 0.262 | -0.25 | 0.51 [0.50 - 0.53] |  |  |
|  |  |  |  | 65 to 74 | 64,515 | 50,110 | 14,405 | 0.223 | -0.29 | 0.44 [0.42 - 0.45] |  |  |
|  |  |  |  | ≥75 | 39,622 | 31,425 | 8,197 | 0.207 | -0.30 | 0.40 [0.39 - 0.42] |  |  |
|  |  |  | Sex | Male | 158,793 | 113,948 | 44,845 | 0.282 | - | 1.00 (reference) |  |  |
|  |  |  |  | Female | 130,845 | 94,569 | 36,276 | 0.277 | -0.01 | 0.98 [0.97 - 0.99] |  |  |
|  |  |  | SEP (NZdep06 quintile) | 5^th^ quintile (low) | 39,766 | 34,289 | 5,477 | 0.138 | - | 1.00 (reference) |  |  |
|  |  |  |  | 4^th^ quintile | 42,291 | 36,115 | 6,176 | 0.146 | 0.01 | 0.87 [0.85 - 0.88] |  |  |
|  |  |  |  | 3^rd^ quintile | 54,267 | 44,100 | 10,167 | 0.187 | 0.05 | 0.75 [0.74 - 0.77] |  |  |
|  |  |  |  | 2^nd^ quintile | 62,381 | 48,946 | 13,435 | 0.215 | 0.08 | 0.59 [0.57 - 0.60] |  |  |
|  |  |  |  | 1^st^ quintile (high) | 71,934 | 54,069 | 17,865 | 0.248 | 0.11 | 0.56 [0.54 - 0.57] |  |  |
|  |  |  |  | Unspecified | 19,027 | 17,779 | 1,248 | 0.066 | -0.07 | 0.26 [ 0.25 - 0.27] |  |  |
|  |  |  | Ethnicity (prioritised) | European | 181,753 | 137,005 | 44,748 | 0.246 | - | 1.00 (reference) |  |  |
|  |  |  |  | Maori | 25,129 | 15,981 | 9,148 | 0.364 | 0.12 | 1.48 [1.45 - 1.51] |  |  |
|  |  |  |  | Pacific Peoples | 16,429 | 9,348 | 7,081 | 0.431 | 0.18 | 1.75 [1.72 - 1.78] |  |  |
|  |  |  |  | Asian | 21,663 | 13,908 | 7,755 | 0.358 | 0.11 | 1.45 [1.43 - 1.48] |  |  |
|  |  |  |  | MELAA | 17,902 | 12,906 | 4,996 | 0.279 | 0.03 | 1.13 [1.11 - 1.16] |  |  |
|  |  |  |  | Other | 113 | 89 | 24 | 0.212 | -0.03 | 0.86 [0.61 - 1.23] |  |  |
|  |  |  |  | Unknown | 26,677 | 19,319 | 7,358 | 0.276 | 0.03 | 1.12 [1.10 - 1.14] |  |  |
| Engebretsen (2024)^f^ | Both | No | Age group | 18 to 54 | 1,207 | - | | | | | Model includes number of statins (≥3 vs. <3) dispensed before PCSK9 inhibitor initiation, region of residence, indication, and initiation year. | 1.00 (reference) |
|  |  |  |  | 55 to 63 | 1,317 |  |  |  |  |  |  | 0.64 [0.51 - 0.81] |
|  |  |  |  | 64 to 70 | 1,188 |  |  |  |  |  |  | 0.64 [0.50 - 0.81] |
|  |  |  |  | 71 to 80 | 1,072 |  |  |  |  |  |  | 0.57 [0.45 - 0.72] |
|  |  |  | Sex | Male | 2,788 |  |  |  |  |  |  | 1.00 (reference) |
|  |  |  |  | Female | 1,996 |  |  |  |  |  |  | 1.63 [1.38 - 1.92] |

^a^All univariate effect estimates are unadjusted risk ratios, except for those from Malo *et al.* (2017) and Chidwick K *et al*. (2018), who reported unadjusted hazard ratios, and Halava *et al*. (2016), who reported unadjusted odds ratios. ^b^All effect estimates from adjusted models are adjusted hazard ratios, except for those from Halava *et al*. (2016) and Sigglekow F *et al*. (2020), who reported adjusted odds ratios. ^c^Chidwick K *et al*. (2018) used IMD quintiles as their SEP measure, but these were in reverse order compared to other papers – quintile 5 was ‘least deprived’ where other authors tend to label this quintile as ‘most deprived’.^26^ ^d^For the Engebretsen *et al*. (2024) hazard ratios, a 180-day treatment gap was used to define discontinuation.^51^ Abbreviations: aHR, adjusted hazard ratio; aOR, adjusted odds ratio; RR, risk ratio; SEP, socioeconomic position.

**Table S8 – Measures of association between sociodemographic categories and antihypertensive discontinuation.**

| **Study or subgroup** | **Prevention** | **Switch as discontinuation?** | **Variable** | **Group** | **Total** | **Persistent** | **Discontinued** | **Proportion discontinued (3 d.p.)** | **Risk difference (2 d.p.)** | **Univariate^a^** | **Multivariate^a^** | |
| --- | --- | --- | --- | --- | --- | --- | --- | --- | --- | --- | --- | --- |
|  |  |  |  |  |  |  |  |  |  | **RR [95% CI]** | **Model description** | **aHR/aOR/aRR [95% CI]** |
| Ah (2015) [Discontinued all AHTs] | Primary | No | Age group | 18 to 64 | 34,931 | - | | | | | Model includes CCI score, insurance type, depression, dementia, dyslipidemia, and initial AHT class. | 1.00 (reference) |
|  |  |  |  | 65 to 79 | 9,131 |  |  |  |  |  |  | 0.94 [0.90 – 0.97] |
|  |  |  |  | ≥80 | 1,725 |  |  |  |  |  |  | 1.14 [1.05 – 1.23] |
|  |  |  | Sex | Male | 23,847 |  |  |  |  |  |  | 1.00 (reference) |
|  |  |  |  | Female | 21,940 |  |  |  |  |  |  | 0.92 [0.89 – 0.95] |
| Ah (2015) [Discontinued index AHT] | Primary | Yes | Age group | 18 to 64 | 34,931 | - | | | | | Model includes CCI score, insurance type, depression, dementia, dyslipidemia, and initial AHT class. | 1.00 (reference) |
|  |  |  |  | 65 to 79 | 9,131 |  |  |  |  |  |  | 0.87 [0.85 – 0.90] |
|  |  |  |  | ≥80 | 1,725 |  |  |  |  |  |  | 0.95 [0.89 - 1.02] |
|  |  |  | Sex | Male | 23,847 |  |  |  |  |  |  | 1.00 (reference) |
|  |  |  |  | Female | 21,940 |  |  |  |  |  |  | 0.88 [0.86 – 0.90] |
| Ah (2016) [Discontinued all AHTs] | Primary | No | Age group | 18 to 64 | 43,954 | - | | | | | Initial ARB type was the main exposure. Model includes insurance type. comorbidity score, depression, dementia, and dyslipidemia. | 1.00 (reference) |
|  |  |  |  | 65 to 79 | 9,833 |  |  |  |  |  |  | 1.04 [1.00 – 1.08] |
|  |  |  |  | ≥80 | 1,717 |  |  |  |  |  |  | 1.38 [1.27 – 1.49]^a^ |
|  |  |  | Sex | Male | 29,350 |  |  |  |  |  |  | 1.00 (reference) |
|  |  |  |  | Female | 26,154 |  |  |  |  |  |  | 0.80 [0.78 – 0.82] |
| Ah (2016)  [Discontinued index ARB] | Primary | Yes | Age group | 18 to 64 | 43,954 | - | | | | | Initial ARB type was the main exposure. Model includes insurance type. comorbidity score, depression, dementia, and dyslipidemia. | 1.00 (reference) |
|  |  |  |  | 65 to 79 | 9,833 |  |  |  |  |  |  | 1.01 [0.98 – 1.04] |
|  |  |  |  | ≥80 | 1,717 |  |  |  |  |  |  | 1.25 [1.17 – 1.34] |
|  |  |  | Sex | Male | 29,350 |  |  |  |  |  |  | 1.00 (reference) |
|  |  |  |  | Female | 26,154 |  |  |  |  |  |  | 0.86 [0.84 - 0.88] |
| Beall (2022)^b^ | Primary | No | Age group | 18 to 45 | 27,933 | 10,895 | 17,038 | 0.610 | - | 1.00 (reference) | - | |
|  |  |  |  | 45 to <65 | 55,911 | 27,547 | 28,364 | 0.507 | -0.10 | 0.83 [0.82 - 0.84] |  |  |
|  |  |  |  | ≥65 | 19,388 | 10,594 | 8,794 | 0.454 | -0.16 | 0.74 [0.73 - 0.76] |  |  |
|  |  |  | Sex | Male | 56,283 | 25,906 | 30,377 | 0.540 | - | 1.00 (reference) |  |  |
|  |  |  |  | Female | 46,949 | 23,130 | 23,819 | 0.507 | -0.03 | 0.94 [0.93 - 0.95] |  |  |
|  |  |  | SEP (Neighbour-hood income quintile) | 1^st^ (low SEP) | 24,465 | 10,846 | 13,619 | 0.557 | - | 1.00 (reference) |  |  |
|  |  |  |  | 2^nd^ | 22,429 | 10,647 | 11,782 | 0.525 | -0.03 | 0.94 [0.93 - 0.96] |  |  |
|  |  |  |  | 3^rd^ | 20,274 | 9,672 | 10,602 | 0.523 | -0.03 | 0.94 [0.92 - 0.96] |  |  |
|  |  |  |  | 4^th^ | 18,409 | 8,901 | 9,508 | 0.516 | -0.04 | 0.93 [0.91 - 0.94] |  |  |
|  |  |  |  | 5^th^ (high SEP) | 17,242 | 8,793 | 8,449 | 0.490 | -0.07 | 0.88 [0.86 - 0.90] |  |  |
|  |  |  |  | Missing | 413 | 177 | 236 | - | | |  |  |
|  |  |  | Ethnicity (proportion of visible minority) | <15% | 30,892 | 15,458 | 15,434 | 0.500 | - | 1.00 (reference) |  |  |
|  |  |  |  | 15 to 29% | 28,302 | 13,755 | 14,547 | 0.514 | 0.01 | 1.03 [1.01 - 1.05] |  |  |
|  |  |  |  | 30 to 49% | 23,985 | 11,145 | 12,840 | 0.535 | 0.04 | 1.07 [1.05 - 1.09] |  |  |
|  |  |  |  | ≥50% | 19,662 | 8,527 | 11,135 | 0.566 | 0.07 | 1.13 [1.11 - 1.15] |  |  |
|  |  |  |  | Missing | 391 | 151 | 240 | - | | |  |  |
| Eastwood (2023)^c^ | Primary | Yes | Age group | 18 to 39 | 13,891 | 11,946 | 1,945 | 0.140 | - | 1.00 (reference) | Models included age, sex, ethnicity, systolic blood pressure, socioeconomic position, CVD risk factors, comorbidity, healthcare usage, polypharmacy, and antihypertensive class, in complete case analyses. The second set of results for condensed ethnic categories are for an analysis performed using the maximum gap method and a 182 day gap to denote discontinuation. All other results used a 90 day gap. | 1.25 [1.05 - 1.52] |
|  |  |  |  | 40 to 49 | 36,730 | 32,322 | 4,408 | 0.120 | -0.02 | 0.86 [0.82 - 0.90] |  | 1.14 [0.99 - 1.32] |
|  |  |  |  | 50 to 59 | 55,254 | 49,729 | 5,525 | 0.100 | -0.04 | 0.71 [0.68 - 0.75] |  | 0.97 [0.88 - 1.08] |
|  |  |  |  | 60 to 69 | 56,611 | 52,082 | 4,529 | 0.080 | -0.06 | 0.57 [0.54 - 0.60] |  | 0.85 [0.80 - 0.92] |
|  |  |  |  | ≥70 | 53,163 | 48,910 | 4,253 | 0.080 | -0.06 | 0.57 [0.54 - 0.60] |  | 1.00 (reference) |
|  |  |  | Sex | Male | 116,034 | 104,431 | 11,603 | 0.100 | - | 1.00 (reference) |  | 1.00 (reference) |
|  |  |  |  | Female | 99,514 | 90,558 | 8,956 | 0.090 | -0.01 | 0.90 [0.88 - 0.92] |  | 0.91 [0.88 – 0.94] |
|  |  |  | SEP (Practice-level IMD quintile) | 5^th^ (low SEP) | 31,397 | 28,571 | 2,826 | 0.090 | - | 1.00 (reference) |  | 1.00 (reference) |
|  |  |  |  | 4^th^ | 36,051 | 32,806 | 3,245 | 0.090 | 0.00 | 1.00 [0.95 - 1.05] |  | 0.99 [0.91 - 1.08] |
|  |  |  |  | 3^rd^ | 41,854 | 38,087 | 3,767 | 0.090 | 0.00 | 1.00 [0.95 - 1.05] |  | 0.99 [0.91 - 1.06] |
|  |  |  |  | 2^nd^ | 46,850 | 42,165 | 4,685 | 0.100 | 0.01 | 1.11 [1.06 - 1.16] |  | 1.04 [0.95 - 1.14] |
|  |  |  |  | 1^st^ (high SEP) | 59,256 | 53,330 | 5,926 | 0.100 | 0.01 | 1.11 [1.06 - 1.16] |  | 1.06 [0.99 - 1.14] |
|  |  |  | Ethnicity (condensed categories) | European | 188,354 | 171,402 | 16,952 | 0.090 | - | 1.00 (reference) |  | 1.00 (reference) |
|  |  |  |  | South Asian | 7,857 | 6,443 | 1,414 | 0.180 | 0.09 | 2.00 [1.90 - 2.10] |  | 2.08 [1.96 - 2.22] |
|  |  |  |  | African/African-Caribbean | 5,482 | 4,221 | 1,261 | 0.230 | 0.14 | 2.56 [2.43 - 2.69] |  | 2.63 [2.38 - 2.86] |
|  |  |  | Ethnicity (expanded categories) | British | 179,439 | - | | | | |  | 1.00 (reference) |
|  |  |  |  | Irish | 1,853 |  |  |  |  |  |  | 1.14 [1.02 - 1.25] |
|  |  |  |  | Other White | 7,203 |  |  |  |  |  |  | 1.72 [1.56 - 1.89] |
|  |  |  |  | Indian | 3,325 |  |  |  |  |  |  | 1.85 [1.69 - 2.00] |
|  |  |  |  | Pakistani | 1,732 |  |  |  |  |  |  | 1.75 [1.54 - 1.96] |
|  |  |  |  | Bangladeshi | 654 |  |  |  |  |  |  | 1.54 [1.28 - 1.82] |
|  |  |  |  | Other South Asian | 2,200 |  |  |  |  |  |  | 1.82 [1.61 - 2.04] |
|  |  |  |  | Caribbean | 1,570 |  |  |  |  |  |  | 1.59 [1.41 - 1.82] |
|  |  |  |  | African | 3,118 |  |  |  |  |  |  | 2.33 [2.13 - 2.56] |
|  |  |  |  | Other Black | 812 |  |  |  |  |  |  | 1.69 [1.49 - 1.92] |
|  |  |  | Ethnicity (condensed categories;  182 day grace period) | European | 188,361 | 180,827 | 7,534 | 0.040 | - | 1.00 (reference) |  | 1.00 (reference) |
|  |  |  |  | South Asian | 7,820 | 7,273 | 547 | 0.070 | 0.03 | 1.75 [1.61 - 1.90] |  | 1.89 [1.69 - 2.08] |
|  |  |  |  | African/African-Caribbean | 5,502 | 4,952 | 550 | 0.100 | 0.06 | 2.50 [2.30 - 2.71] |  | 2.27 [2.04 - 2.50] |
| Muntner (2023) | Both | Yes | Age group | 20 to 44 | 253,181 | - | | | | | Model included the calendar year of initiation age, sex, index drug class, number of AHTs being taken, mode of prescription fill, diabetes, CVD, heart failure, CKD, depression, serious fall injury, and polypharmacy. | 1.00 (reference) |
|  |  |  |  | 45 to 64 | 2,197,303 |  |  |  |  |  |  | 0.78 [0.77 - 0.79] |
|  |  |  |  | 65 to 79 | 500,655 |  |  |  |  |  |  | 0.63 [0.62 - 0.64] |
|  |  |  |  | ≥80 | 182,914 |  |  |  |  |  |  | 0.76 [0.75 - 0.78] |
|  |  |  | Sex | Male | 1,689,699 |  |  |  |  |  |  | 1.00 (reference) |
|  |  |  |  | Female | 1,444,354 |  |  |  |  |  |  | 1.03 [1.02 – 1.04] |

^a^All univariate effect estimates are unadjusted risk ratios. All multivariate effect estimates are aHRs, except for those from Eastwood SV *et al*. (2023) which reported aORs and Muntner P *et al*. (2023) which reported aRRs.^48,50^ ^b^For Ah Y *et al*. (2016), the aHR [95% CI] for discontinuation of all antihypertensives is reported in the text as 1.38 [1.27 - 1.59], which is asymmetric.^14^ We assumed this was a copying error and assumed an upper 95% CI bound of 1.49 instead, which produces a symmetrical CI. ^c^For Beall RF *et al*. (2022), a 7 day treatment gap was used to define discontinuation.^45 d^For Eastwood SV *et al.* (2023), the number of participants used for our unadjusted analyses were calculated by multiplying out proportions from Supplementary Table 11 of that paper.^48^ The counts are therefore non-exact. The adjusted analysis in Eastwood SV *et al.* (2023) was computed using an overall sample of 201,179 individuals. Abbreviations: aHR, adjusted hazard ratio; AHTs, antihypertensives; aOR, adjusted odds ratio; ARBs, angiotensin-receptor blockers; aRR, adjusted risk ratio; CCI, Charlson comorbidity index; CKD, chronic kidney disease; CVD, cardiovascular disease; LLTs, lipid-lowering therapies; SEP, socioeconomic position.

**Figure S1 – Association of age group with discontinuation of (A) lipid-lowering therapies and (B) antihypertensives.**

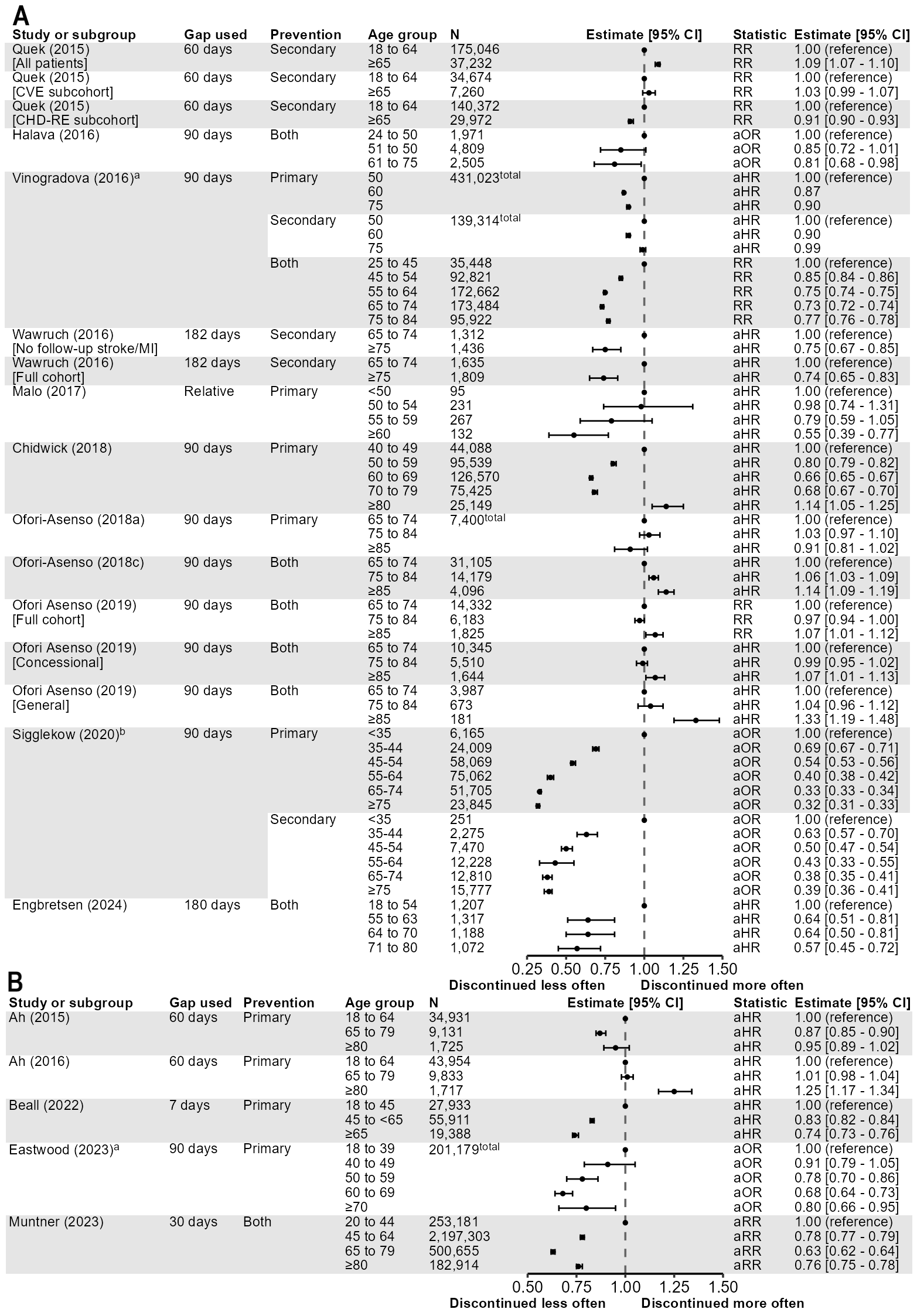

^a^Vinogradova *et al*. (2016) results for primary and secondary CVD prevention, Eastwood *et al*. (2023) results, and Sigglekow *et al*. (2020) results include plotted estimates/95% CIs where the reference age was changed from from 60 to 50, the reference age group was changed from ‘≥70’ to ‘18 to 39’, and the reference age group was changed from ’55 to 64’ to ‘<35’, respectively.^22,41,48^
